# Adapting an intervention to improve hypertension care for adults with HIV in Tanzania: Co-design of the Community Health Worker Optimization of Antihypertensive Care in HIV (COACH) intervention

**DOI:** 10.1101/2025.11.26.25340978

**Authors:** Preeti Manavalan, Blandina T. Mmbaga, Nathan M. Thielman, Melissa H. Watt, Spencer F. Sumner, Tazeen H. Jafar, Hayden B. Bosworth, Francis M. Sakita, Lisa Wanda, Kelvin F. Haukila, Godfrey Kweka, Jerome Mlangi, Pankrasi Shayo, Julian T. Hertz

## Abstract

**Introduction:** There is a large burden of uncontrolled hypertension among people with HIV (PWH) in sub-Saharan Africa (SSA), including in Tanzania. Yet, few evidence-based interventions to improve hypertension control have been adapted for use in PWH in this region. This study describes the adaptation process of an evidence-based hypertension intervention to develop the *Community Health Worker Optimization of Antihypertensive Care in HIV* (*COACH*) intervention, a multi-component strategy designed to improve blood pressure control among Tanzanians with HIV and hypertension.

**Methods:** A 27 member interdisciplinary intervention design team consisting of HIV and hypertension clinicians, nurses, community health workers (CHWs), pharmacists, social workers and patients with HIV and hypertension from Tanzania met biweekly from May 2024 to October 2024. The design team used the Assessment-Decision-Adaptation-Production-Topical Experts-Integration-Training-Testing (ADAPT-ITT) framework supported by participatory co-design principles to iteratively adapt the intervention to the local context.

**Results:** To address the unique needs of PWH and hypertension in Tanzania, we iteratively adapted an evidence-based CHW intervention for hypertension care originally developed in Asia (*Control of Blood Pressure and Risk Attenuation—COBRA*), resulting in development of the *COACH* intervention for the HIV clinical setting in Tanzania. *COACH*, includes five key components: 1) CHW-delivered hypertension counselling integrated into HIV clinic visits, 2) Integration of routine blood pressure monitoring and referrals for antihypertensive medication management in the HIV clinic, 3) Hypertension management training for HIV providers and creation of an antihypertensive treatment algorithm, 4) CHW care navigation and coordination of hypertension care in the HIV clinic, and 5) Subsidization of antihypertensive medications.

**Conclusions:** *COACH* is one of the first contextually-tailored interventions developed to address hypertension care among PWH in Tanzania. A pilot feasibility study of the intervention is in process and future studies will evaluate the implementation and clinical effectiveness outcomes of the *COACH* intervention. The rigorous, systematic application of the ADAPT-ITT framework to iteratively develop *COACH* supports reproducibility of the adaptation process, and strengthens the potential for *COACH* core components to be highly relevant for PWH with hypertension in other resource limited settings worldwide.

## Introduction

People with HIV (PWH) face increased risk for cardiovascular disease (CVD), and the burden of CVD is growing in sub-Saharan Africa (SSA) [1–4]. Hypertension is the leading risk for CVD morbidity and death [5], yet its management among PWH in SSA remains frequently suboptimal [6]. Our previous work in Tanzania found that most PWH and hypertension were not receiving antihypertensive treatment and almost all had uncontrolled blood pressure, portending an increased risk of CVD-related death [7–11]. Thus, there is an urgent need for implementation of evidence-based interventions (EBIs) to address the rising burden of hypertension and CVD risk in Tanzania and other similar settings. Yet, few EBIs to improve hypertension control have been adapted for use in PWH in this region [12, 13].

Hypertension interventions that are culturally and contextually derived are needed to maximize impact and effectiveness in real-world settings [14]. We previously developed a *community health worker (CHW)-delivered Hypertension Management Pilot* (*CHAMP*) to improve hypertension care among PWH in northern Tanzania. In a small pilot feasibility study in one HIV clinic, we found CHAMP to be highly feasible and acceptable [15]. However, while we noted substantial improvements in hypertension care engagement and an 18-point reduction in median systolic blood pressure, *CHAMP* was only piloted among a small number of participants over a short duration of time and was not powered to test effectiveness [15]. Evidence-based hypertension interventions in other regions have demonstrated clinical efficacy and reduced all-cause mortality. For example, *Control of Blood Pressure and Risk Attenuation* (*COBRA*), is a community-based multi-component intervention consisting of CHW-delivered education and blood pressure monitoring in the community with protocolized referrals to physicians. *COBRA* demonstrated efficacy in blood pressure control and in reducing all-cause mortality in a cluster randomized trial across 3 Southeast Asian countries [16–19]. However, *COBRA* has not been studied among PWH nor in SSA. Adaptation of EBIs like *COBRA*, that are tailored to the HIV clinical setting, have the potential to bridge the gaps in hypertension care among PWH in Tanzania.

Development and adaptation of EBIs, especially across diverse clinical contexts, requires grounding in theoretical models using implementation science frameworks to ensure intervention feasibility, acceptability and scale-up [20, 21]. ADAPT-ITT (Assessment-Decision-Adaptation-Production-Topical Experts-Integration-Training-Testing) is a pragmatic framework utilizing iterative, experiential processes to adapt and implement EBIs in a diversity of settings and contexts [22]. Additionally, participatory co-design is a collaborative process that involves community members, researchers and other stakeholders in the design process of interventions and is based on the principle that communities and community members are experts [23–26]. Use of the ADAPT-ITT framework and participatory co-design ensures that those who would benefit most from an intervention are involved in its adaptation, thus meeting local needs for hypertension care.

In this study, we used the ADAPT-ITT framework, supported by a participatory co-design approach, to iteratively adapt the evidence-based *COBRA* intervention, integrating lessons from our previous pilot work in Tanzania, to develop the *Community Health Worker Optimization of Antihypertensive Care in HIV* (*COACH*) intervention to improve hypertension care and clinical outcomes among PWH in Tanzania. We present results from the adaptation process and the components of the final *COACH* intervention.

## Methods

### Setting

This study was conducted at two publicly funded HIV clinics in northern Tanzania: Majengo Care and Treatment Center and Pasua Care and Treatment Center. With support from the Tanzanian Ministry of Health and the Elizabeth Glaser Pediatric AIDS Foundation (EGPAF), the Majengo and Pasua Care and Treatment Centers provide free HIV care to over 2300 adults (approximately 75% women) with HIV. Our previous work identified that 20-35% of patients enrolled in HIV care in this region were hypertensive [7, 8]. While both HIV clinic sites provide antiretroviral therapy (ART) at no cost, hypertension screening and management has not been routinized at either clinic. Both clinics are staffed by clinical officers (mid-level advanced practice providers) trained in the provision of HIV care and services. Similar to other clinical settings in Tanzania, PWH with hypertension typically receive hypertension care with a separate provider at a separate clinic and usually pay out of pocket for these hypertension services and treatment.

### Intervention Design Team

An Intervention design team composed of key stakeholder groups was assembled to adapt the *COBRA* intervention for the local context. The intervention design team consisted of 27 members who were purposively selected to allow for diversity in expertise, including physicians and clinical officers with substantive experience treating hypertension and HIV, pharmacists, nurses, CHWs, social workers, patients with HIV and hypertension, and local and national public health administrators (Table 1).

**Table 1.** Characteristics of Intervention Design Team Members, 2024 (n=27)

| Characteristic | n (%) |
| --- | --- |
| Sex |  |
| Female | 15 (55.5%) |
| Male | 12 (44.5%) |
| Area of expertise* |  |
| HIV provider | 5 (18.5%) |
| Patient with HIV and hypertension | 5 (18.5%) |
| Nurse | 5 (18.5%) |
| Hypertension provider | 4 (14.8%) |
| CHW | 3 (11.2%) |
| Pharmacist | 2 (7.4%) |
| Social worker | 2 (7.4%) |
| Regional public health administrator | 2 (7.4%) |
| National public health administrator | 1 (3.7%) |

\*Areas of expertise are not mutually exclusive

### Study overview

The formative data presented here reflect the process of developing the *COACH* intervention through adaptation of an EBI (*COBRA*) and integration of lessons learned from our previous *CHAMP* intervention in Tanzania. The intervention was developed iteratively through a series of 14 intervention design team meetings which occurred approximately biweekly between May 2024 and October 2024, and the adaptation process was guided by the ADAPT-ITT framework. Meetings occurred in-person at one of the study sites, however, a hybrid format was used to allow some design team and research study members to participate virtually. Meetings were moderated by a member of the research team, typically lasted for 2 hours, and members were provided 20,000 Tanzanian Shillings ($8 USD) per meeting for compensation for their time and travel. There was a high level of engagement at each meeting, with attendance among intervention design team members ranging between 81-100% per meeting. Meetings were conducted in Swahili, with real-time translation to English for non-Swahili speaking members of the research team. Minutes summarizing the discussion were sent to all members of the research and intervention design teams at the end of each meeting for their review and for additional feedback. Assembly of the intervention design team to assist in intervention adaptation was based on participatory co-design principles and allowed for individuals who would benefit the most from the intervention to be actively involved in intervention development, thus meeting the local needs for hypertension care. The overall goals of the *COACH* intervention were to: improve hypertension control among PWH, improve hypertension awareness and knowledge among PWH and their providers, and improve hypertension care engagement and antihypertensive adherence among PWH.

### Intervention development process guided by ADAPT-ITT

The ADAPT-ITT framework has eight steps: 1) *assessment* to understand the target population and the capacity of the organization to implement the intervention; 2) *decision* to adopt or adapt an intervention; 3) *adaptation* through participatory design; 4) *production* of the adapted EBI; 5) consulting *topical experts*; 6) *integrating* feedback into the adapted EBI; 7) *training* personnel involved in the intervention; and 8) *testing* the adapted EBI [22].

#### Assessment

The *COACH* intervention was informed by our previously collected qualitative and quantitative data regarding barriers and facilitators of hypertension care for PWH in Tanzania as well as our data generated from the *CHAMP* pilot study [7–11, 15, 27, 28]. The results from this formative work highlighted the need for integrated HIV and hypertension care, improved hypertension education and awareness among patients and providers, and the potential for task-shifting approaches to bridge the gaps in hypertension care for PWH. These findings have been described elsewhere and are also summarized Table 2. The study team presented the results to intervention design team members, and in turn, the members provided input on the intervention in light of these data and in consideration of current Tanzanian policies and infrastructure.

**Table 2.**
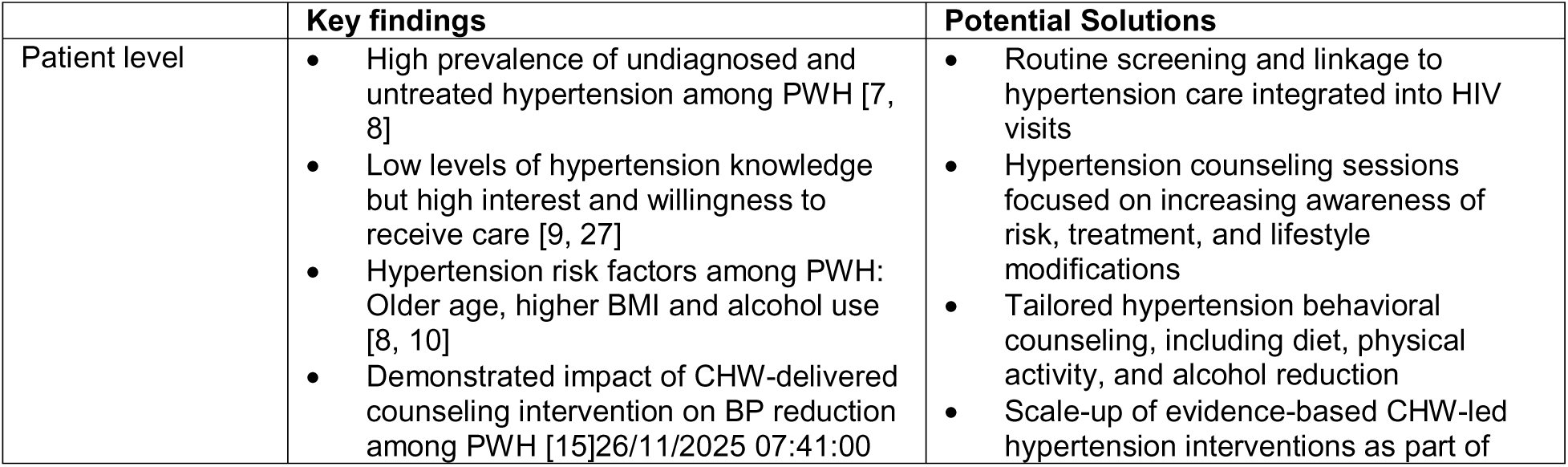
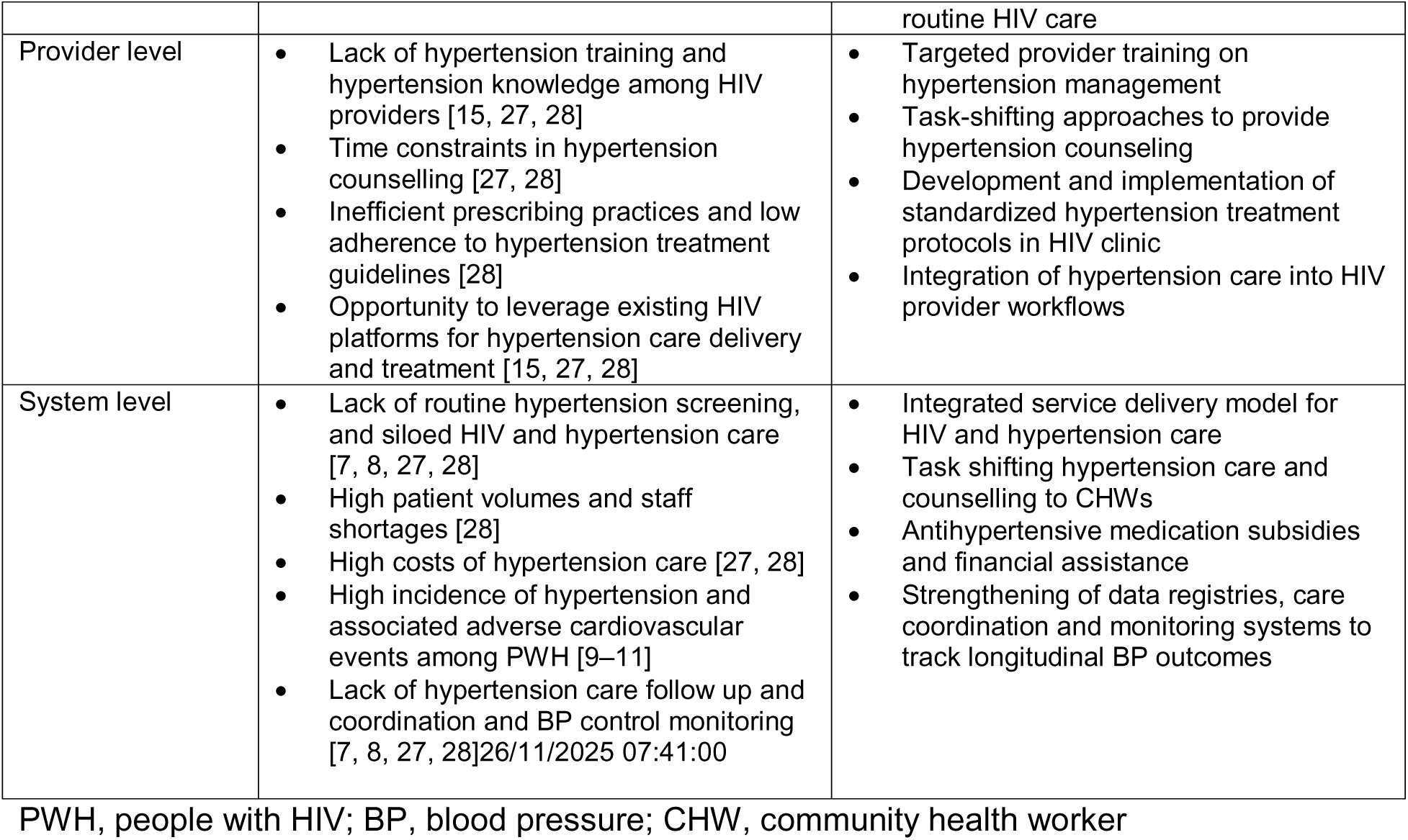
Summary of local research findings informing *COACH* intervention solutions by patient, provider and system level.

#### Decision

In the Decision phase, we considered data from the Assessment phase to decide whether it would be appropriate to adapt the evidence-based *COBRA* intervention by incorporating key components from the *CHAMP* pilot. *COBRA* effectively improved blood pressure control in multiple low-and middle-income countries in Southeast Asia by utilizing behavior change principles and addressing gaps in primary care due to paucity of physicians, lack of patient education and provider training, and high costs of hypertension care. *COBRA* included 5 core components: 1) CHW-home based hypertension counselling, 2) CHW measuring and monitoring of home blood pressure and referral to a local clinician if elevated, 3) hypertension training for clinic providers and creation of an evidence-based algorithm for antihypertensive medication treatment, 4) a clinic nurse coordinator who tracked all hypertension referrals, and 5) low-cost antihypertensive treatment for patients. Of note, while *COBRA* intended to include free travel vouchers to the clinic and free antihypertensive medications as part of their intervention [29], this was not implemented, and instead, the cost of medications and travel were borne primarily by the patients and low-cost antihypertensive prescriptions were encouraged in the clinics [16]. We aimed to integrate key elements of the *CHAMP* pilot into these 5 *COBRA* components with the goal of adapting the intervention for delivery within the Tanzanian HIV clinical setting (Figure 1). Decisions took into consideration the feasibility and future scalability of the adapted intervention given limited resources in the hypertension clinic and high volume of patients in the HIV clinic. Therefore, we sought to make use of existing resources in the HIV clinic. The intervention design team proposed this would be a scalable approach as it could easily be administered to patients already receiving HIV care and would leverage existing HIV clinic resources and staff including CHWs, providers and pharmacists for delivery of intervention components. The adapted intervention was designed with the long-term goal of integration into existing HIV clinic appointments.

**Figure 1.**
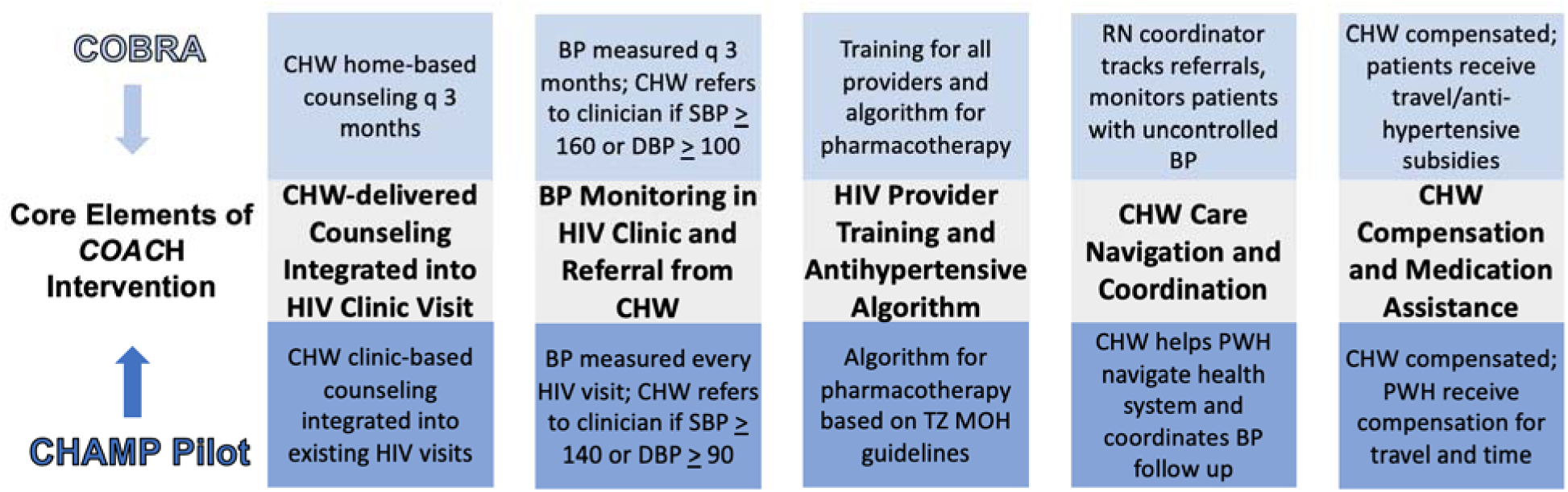
Overlap of the core components of COBRA and CHAMP integrated and adapted in COACH.

#### Adaptation

Between May 2024 and October 2024, the intervention design team met approximately biweekly to iteratively adapt the selected hypertension intervention for the HIV clinic setting in Tanzania. During meetings, design team members “workshopped” the original *COBRA* and *CHAMP* material from existing protocols and manuscripts. Through group discussions and consensus, members proposed adaptations, identified emerging content, and developed the initial framework for the *COACH* intervention.

#### Production, Topical Experts and Integration

As a next step, draft materials of all intervention components were then created by the study team and were shared with intervention design team members to elicit feedback. Additional input from external U.S. and Tanzanian topical experts in implementation science, social science, HIV and hypertension was also sought for further refinement of draft materials. In an iterative cycle, feedback from members and topical experts were integrated into the intervention materials until creation of the adapted intervention product.

#### Testing and Training

The adapted intervention was then pre-tested with 5 purposively selected patients with lived experience of HIV and hypertension to obtain qualitative feedback and explore potential challenges and solutions to further refine and finalize the intervention content and delivery processes.

After finalizing the adapted intervention, the research team developed a protocol to train CHWs employed at the HIV clinic sites in the delivery of the *COACH* intervention sessions. CHWs received a series of five training sessions (5 hours each) over the course of 5 days on the finalized *COACH* blood pressure monitoring and counselling curriculum. CHWs were compensated 30,000 TSH ($12 USD) per training session for their time. CHW training sessions were led by members of the study team and training materials focused on appropriate techniques for BP measurement; general knowledge about the risk factors, causes, symptoms, diagnosis and treatment of hypertension; and in-depth review of the finalized *COACH* hypertension counselling curriculum as well as the referral and tracking documents for hypertension care visits with providers. Additionally, as part of the intervention, training sessions were held with HIV providers regarding the hypertension treatment and use of the medication algorithm.

### Ethics

The study received ethical approval from institutional review boards at Duke Health (Pro00090902), KCMC (No. 1454), and the Tanzania National Institute of Medical Research (NIMR/HQ/R.8a/Vol.IX/4615).

## Results

### Results from the ADAPT-ITT process

The process of adapting *COBRA* to include key components of *CHAMP* elicited constructive feedback and identified several key insights related to challenges in the current standard of hypertension care for PWH including hypertension screening, diagnosis, treatment and follow up (see Table 3). Hypertension services were completely siloed from HIV care, hypertension care and treatment was quite costly, and counselling on hypertension treatment adherence was limited due to challenges in the existing infrastructure, including provider shortages, time constraints and lack of training. As such, the integration of *CHAMP* elements into *COBRA* to develop the *COACH* intervention was guided by a strategic focus of overcoming these key challenges. This included integrating hypertension and HIV care, reducing patient costs, and enhancing treatment adherence through a multifaceted approach with an emphasis on task-shifting with CHW-delivered hypertension counselling and care coordination as well as strengthening provider training to optimize patient outcomes.

**Table 3.**
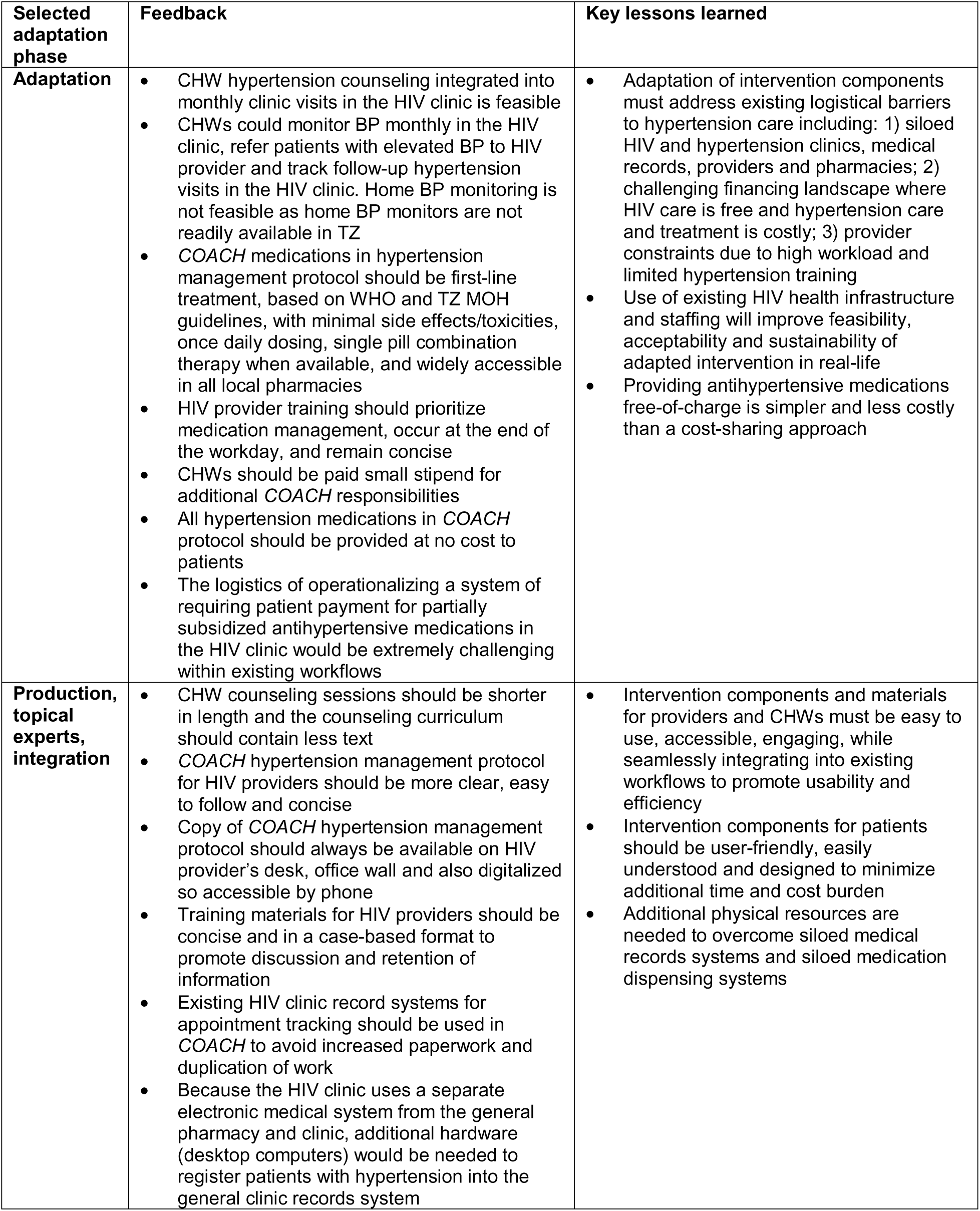
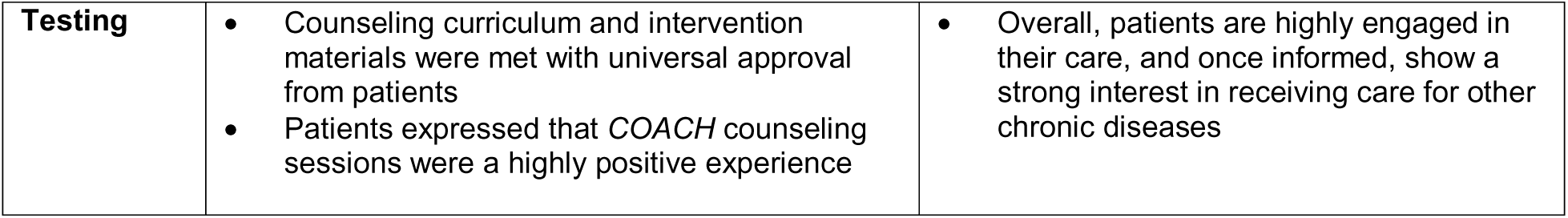
Summary of feedback and key lessons learned from selected components of adaptation process.

As a next step, a draft of the adapted intervention materials was created by the study team and shared with all intervention design team members as well as physician scientists with expertise in the delivery of hypertension care for PWH in resource-limited settings to elicit feedback for further refinement of content. Recommendations for additional refinement focused on condensing, simplifying and streamlining materials for efficiency, ease of use and increased understanding among patients, CHWs and providers. Feedback was integrated into revised draft materials that were once again shared with design team members and topical experts who agreed with the revised materials and confirmed acceptability.

Research staff members next pre-tested all six of the in-person COACH counseling sessions and the telephone sessions with 5 purposively selected patients who received HIV care at one of the two clinic study sites and were also previously diagnosed with hypertension. The counseling session content and materials resonated with the patients and pre-testing confirmed intervention feasibility and acceptability of the intervention materials among all 5 patients. Pretesting helped finalize each counseling session and informed the training protocol materials for CHW delivery of the intervention.

### Resulting COACH intervention

We present the final components of the *COACH* intervention resulting from the intervention adaptation process. Components of the adapted intervention are available in the Supplementary Material (see Additional File 1).

#### Overview of COACH intervention

The *COACH* intervention was designed for PWH with hypertension and for delivery over a 6-month period (see Figure 2). Patients enrolled in HIV care in northern Tanzania typically attend the HIV clinic for HIV care and receipt of ART approximately every 6 months (ranging anywhere between 1 to 6 months), but patients with hypertension seen in the general medical clinic typically are seen by providers on a monthly basis. *COACH* intervention hypertension care visits will occur monthly in the HIV clinic and any existing HIV visit will be integrated with the hypertension visit. Patients who meet study criteria for hypertension will attend monthly in-person sessions at the HIV clinic and will meet with both the CHW and the HIV provider for hypertension counselling and treatment. Participants will also have telephone sessions with a CHW in between the in-person sessions. The CHW will be an existing health worker in the Tanzanian Community-Based HIV Services (CBHS) program. The CBHS program consists of a national cohort of volunteer CHWs who support HIV care engagement in HIV clinics. These individuals routinely provide HIV care counselling in the HIV clinic and perform home visits and outreach in the community for patients who are lost to follow up, and they receive a small monthly stipend from the government for their work. At the time of this study, there were 12 such CHWs at the first HIV clinic site and 13 CHWs at the second clinic site in the CBHS program and four CHWs (two from each site) were selected as CHWs for the *COACH* intervention. All HIV providers at the study sites will participate in the *COACH* intervention for provision of medical management of hypertension.

**Figure 2.**
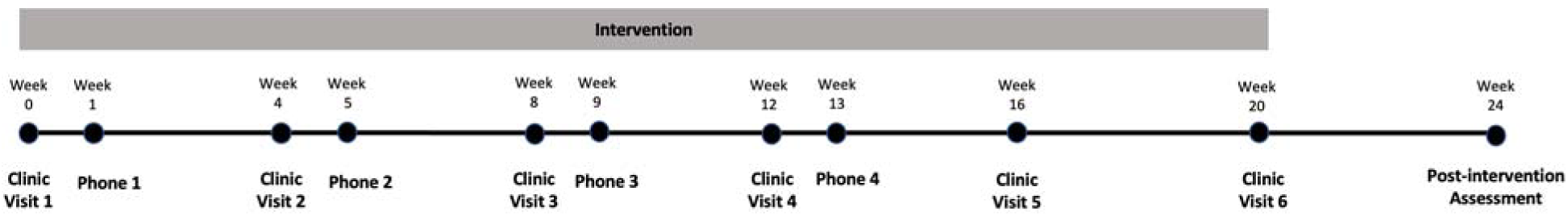
Overview of COACH intervention integrated into HIV clinics in Northern Tanzania.

The resulting *COACH* intervention for PWH and hypertension consists of 5 components. Table 4 summarizes the 5 key components which include: 1) Six CHW-delivered hypertension education counselling sessions integrated into monthly clinic visits in the HIV clinic and four follow-up telephone sessions in between the in-person sessions, 2) Monitoring of blood pressure by the CHW during monthly HIV visits and CHW facilitated referrals to the HIV clinician for antihypertensive management, 3) Training for HIV providers on hypertension management and use of the *COACH* Hypertension Management Protocol, 4) CHW care coordination and navigation for hypertension follow up visits in the HIV clinic, and 5) Subsidization of antihypertensive medications and stipends for the CHWs.

**Table 4.** Components of the COACH intervention for integrating hypertension and HIV care in Tanzania.

| Component | Details |
| --- | --- |
| 1. CHW Counseling | • Six one-on-one monthly counseling sessions between clinic-based CHW and patient in the HIV clinic<br>• Each session lasts for approximately 30 minutes and is guided by a curriculum with hypertension educational content (see Supplementary Material)<br>• Brief, telephone check-in sessions between in-person session focused primarily on antihypertensive medication adherence, troubleshooting issues, and answering additional questions<br>• Patient receives hypertension educational handout |
| 2. Monitoring and Referrals | • CHW measures patient's BP on arrival to the clinic at each monthly visit in the HIV clinic<br>• CHW refers patient to provider in the HIV clinic for initiation or modification of antihypertensive therapy if BP is elevated (SBP ≥ 140 mmHg or DBP ≥ 90 mmHg)<br>• Referrals for antihypertensive care are made to patient's regular HIV provider, rather than to a separate provider |
| 3. Provider Training and Antihypertensive Algorithm | • All HIV clinicians receive initial training session led by a hypertension specialist and a subsequent refresher training<br>• Training sessions cover hypertension diagnosis and management, as well as common treatment misconceptions<br>• Providers follow a standardized hypertension treatment algorithm based on WHO guidelines, with stepwise instructions for titration of therapy (see Supplementary Material) |
| 4. CHW Care Navigation and Coordination | • CHWs maintain a registry of patients with HIV and hypertension<br>• CHWs monitor patient attendance at monthly appointments and contact patients who miss appointments via telephone or home visits to re-schedule their visit<br>• Antihypertensives dispensed directly from the HIV clinic pharmacy, rather than from a separate pharmacy<br>• Patients with HIV and hypertension are registered in the general clinic electronic medical record system to allow for integration of their hypertension care into existing medical records system |
| 5. Subsidies | • Antihypertensives in the treatment algorithm (amlodipine, losartan, and combined losartan/hydrochlorothiazide) provided for free to participants<br>• CHWs receive a monthly supplemental stipend for providing hypertension counseling and care coordination |

CHW, community health worker; BP, blood pressure; SBP, systolic blood pressure; DBP, diastolic blood pressure

#### Five core components of the COACH intervention

##### 1. CHW-delivered hypertension counselling integrated into HIV clinic visits

A total of six in-person CHW-counseling sessions and four telephone sessions were developed. A detailed educational curriculum with accompanying visual aids were created for each session, based on the constructs of the Health Belief Model (see Supplement) [30]. Curriculum content was informed by the CHAMP study and content was confirmed during iterative adaptation with the intervention design team. Each in-person CHW-counseling session was designed to be approximately 20-30 minutes in length and telephone sessions approximately 5-10 minutes in length.

Counseling session 1 focuses on hypertension education including an overview of risk factors, diagnosis, symptoms and complications, and an introduction to treatment of hypertension. The goal of this session is to increase overall awareness to motivate the patient into action.

Counseling session 2 occurs one month after the first in-person session and focuses on the importance of antihypertensive treatment and medication adherence. Counseling session 3 is delivered one month after the second in-person session and focuses on lifestyle modification counselling on healthy diet, weight, exercise, reduction in alcohol use and smoking cessation and creation of an action plan to provide participants with the tools needed to promote behavioral change. Counseling session 4 occurs the following month and highlights the importance of hypertension treatment with a focus on both pharmacologic and non-pharmacologic treatment adherence. Counseling session 5 is delivered one month after session 4 and also focuses on the importance of treatment adherence. The goal of this session is to start to establish a commitment to long-term hypertension treatment. Counseling session 6 occurs one month later and is the final session and the goal is to create long-term commitment to behavioral change and medication use. Telephone sessions are delivered 1 week after the first four in-person sessions and serve as a check in call to answer questions regarding hypertension care and medications, remind the participants of their next in-person appointment and to help build a therapeutic relationship between the participant and the CHW.

##### 2. CHW-facilitated blood pressure monitoring and referrals to HIV clinician for hypertension treatment

At the beginning of every in-person counselling session the CHW measures and records the blood pressure. Following completion of each in-person counseling session, the CHW then refers and accompanies the participant to the HIV clinician providing the recorded BP value so that the clinician can determine the appropriate next steps of antihypertensive treatment with initiation of medication or dose adjustment of current therapy.

##### 3. Creation of an antihypertensive medication management protocol and hypertension management training for HIV providers

A COACH hypertension management protocol, shown in Figure 3, was created based on WHO hypertension treatment guidelines,[31] availability of medications in Tanzania, and feedback from intervention design team members regarding accessibility and cost. Design team members and topical experts favored use of medications with once daily dosing, single pill combination therapy when available, easily tolerated with minimal side effects, and affordable and widely accessible.

**Figure 3.**
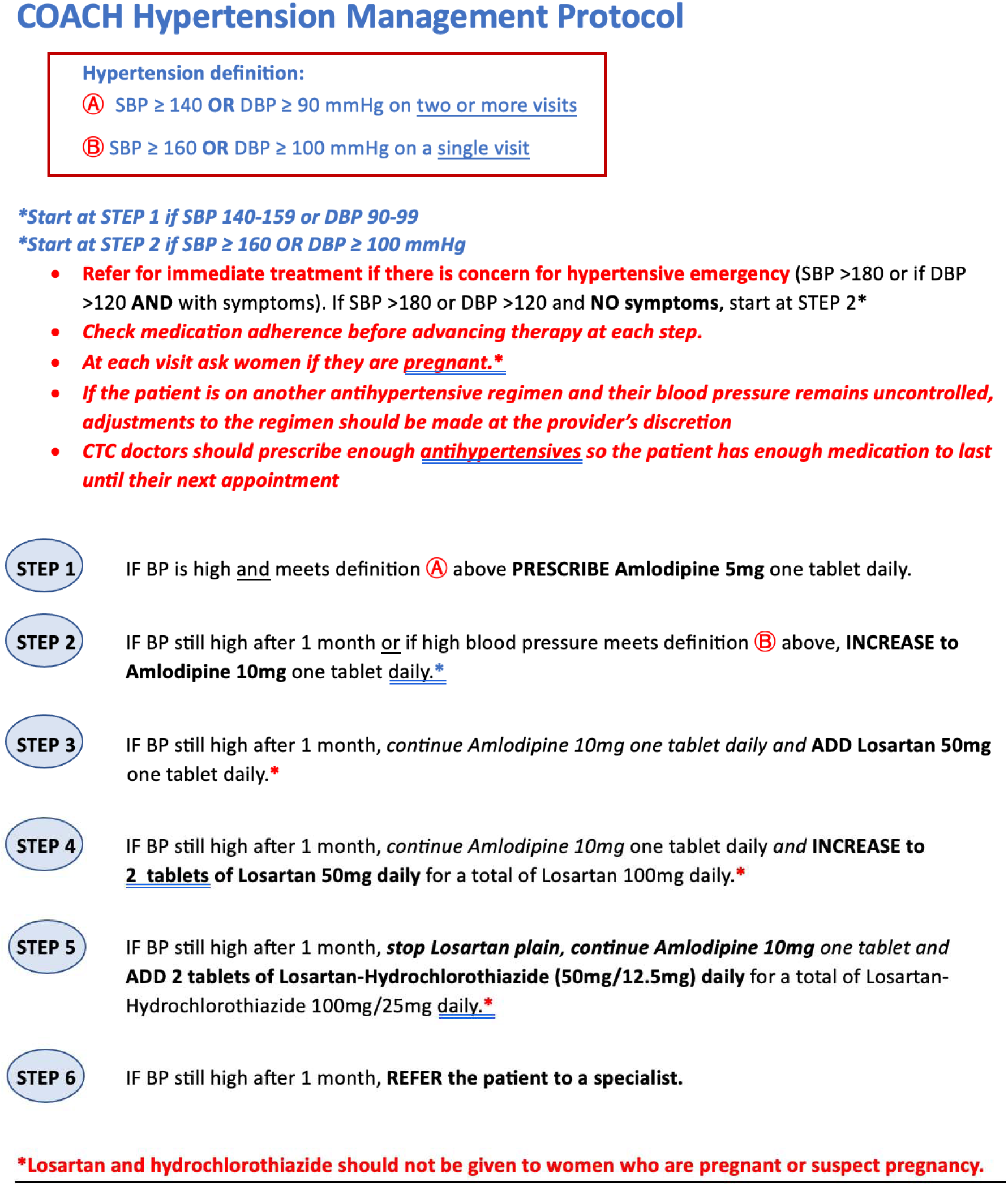
Final *COACH* hypertension management protocol for HIV providers.

HIV clinicians received an initial training session over the course of 2 days (2.5 hours of training per day) on the pharmaceutical management of hypertension with a focus on use of the finalized *COACH* hypertension management protocol that was led by one of the study PIs who has clinical and research expertise in HIV and hypertension and a Tanzanian physician with expertise in hypertension care. A second refresher training session was conducted 4 months later and led by a Tanzanian physician. Trainings emphasized real-life cases and used case-based discussion in small groups to facilitate learner engagement and confirm understanding and mastery of the protocol (see Supplement). Training sessions were held at the end of the work day, and providers were offered tea and snacks and compensated 40,000 TSH ($16 USD) for their time. Engagement during training sessions was high; 100% of HIV providers at both clinic sites attended all training sessions.

##### 4. CHW care navigation and coordination of hypertension care in the HIV clinic

CHWs will maintain a registry of all participants with HIV and hypertension in order to schedule in-person and telephone follow up appointments and track attendance of appointments.

Participants will also receive a telephone call from the CHW 1-2 days prior to their in-person counselling sessions as an appointment reminder. CHWs will utilize the existing HIV clinic protocol for missed HIV appointments for any missed in-person *COACH* session appointments: CHWs will first call participants who missed their scheduled appointment. If unable to get in touch with the participant by phone after three attempts, they will then visit the participant at their home. Additionally, all participants will be registered into the general clinic electronic medical record system so that HIV clinicians and pharmacists will be able to record and monitor antihypertensive medication prescriptions. Antihypertensive medications will be dispensed directly from the HIV clinic pharmacy located within the HIV clinic to optimize integration of care.

##### 5. Subsidization of antihypertensive medications and support services

All antihypertensive medications included in the COACH hypertension management protocol (amlodipine, losartan and combination losartan/HCTZ) will be provided at the HIV pharmacy at no cost to participants for the duration of the study. In addition, each CHW will receive an additional monthly stipend of 120,000 TSH ($48 USD) for providing hypertension counselling, blood pressure monitoring and care coordination.

The final COACH intervention was implemented at both HIV clinic sites beginning in December 2024 as part of a pilot feasibility study. The results of this ongoing pilot trial will be reported separately.

## Discussion

This study reports the adaptation process and the resulting final components of the *Community Health Worker Optimization of Antihypertensive Care in HIV (COACH)* intervention, one of the first systematically-derived and locally tailored interventions to improve hypertension care among PWH in Tanzania. As the burden of CVD continues to rise among the aging population of PWH in SSA [2, 32, 33], contextually-tailored interventions that address hypertension, the leading risk factor for CVD-related mortality [34], are urgently needed. The application of implementation science frameworks in the design of such interventions is critical to ensuring their real-world effectiveness [20]. In this study, we applied participatory co-design principles and utilized the ADAPT-ITT framework to adapt an EBI to ensure its acceptability, appropriateness and sustainability within the local context. Future studies will evaluate implementation and effectiveness of the adapted intervention [35].

To our knowledge, this is one of the first studies from the region in which an evidence-based hypertension intervention was adapted for PWH using an extensive stakeholder participatory approach informed by implementation science methods [36–41]. By systematically applying a rigorous adaptation process, we identified gaps between knowledge and action enabling prioritization of impactful, feasible solutions with high likelihood of sustainability—even in a resource-limited setting [42]. For example, we identified high patient volumes, workforce shortages and siloed and vertical health systems which informed the direction of our intervention [27, 28]. Guided by an interdisciplinary team of community and clinical experts at every step of our iterative process, we believe we have developed an intervention that is both contextually relevant and highly responsive to the hypertension care needs of a priority population of PWH in Tanzania.

*COACH* is a multi-component hypertension intervention that leverages existing HIV infrastructure, task-shifting and an integrated model of care, strategies that address the identified gaps in hypertension care in Tanzania. Substantial investments and responses to the HIV epidemic in SSA have strengthened the HIV health system establishing it as the region’s first large chronic disease program [43–45]. HIV clinics in SSA now represent a valuable opportunity to expand chronic disease management beyond HIV and integrate non-communicable disease (NCD) care, including hypertension treatment [46–48]. Numerous other studies in the region have demonstrated the effectiveness of leveraging existing HIV infrastructure to improve NCD service delivery for PWH [12, 49–51]. Additionally, task shifting to CHWs, who are already embedded within the HIV care model in SSA and routinely provide HIV counseling, offers a sustainable approach to hypertension management [52]. Several other studies have shown improved clinical outcomes when NCD care is task-shifted to CHWs [53, 54]. By incorporating task-shifting and capitalizing on the existing HIV infrastructure to address key barriers to hypertension care for PWH in Tanzania, the *COACH* intervention holds significant promise for effectively bridging the current gaps in care.

Another important insight gained from our study, was that early and frequent engagement with key stakeholders in the intervention design team – including community members with lived experience and healthcare workers with expertise in caring for the target population – facilitated the adaptation of the intervention to the local context. The intentional use of participatory co-design strategies ensured that stakeholder insights were meaningfully incorporated [55, 56]. Nearly all components of the original evidence-based *COBRA* intervention required modification for adaptation in our setting – and intervention design team members focused on adapting the components to address critical logistical barriers to hypertension care faced by PWH in Tanzania while also enhancing usability and efficiency amongst both providers and patients in the HIV clinic.

A limitation of this study was its focus on two public health facilities within a single region in in northern Tanzania. While this allowed for extensive tailoring to a specific context, the focus on a specific region may restrict the applicability of the intervention to other settings. Although additional refinement of the *COACH* intervention may be required prior to its implementation in other settings, our rigorous and systematic application of the evidence-based ADAPT-ITT framework enhances the reproducibility of our adaptation process and strengthens the potential for *COACH* core components to be highly relevant for PWH and hypertension in other settings beyond Tanzania. Additionally, barriers to hypertension care are very similar across resource limited settings in both LMICs as well as high income countries [57]. As such, the *COACH* intervention has potential to be broadly applicable in a plurality of settings and contexts.

## Conclusions

This paper described the development of an intervention to integrate hypertension care into the HIV clinical care in Tanzania to improve hypertension outcomes and reduce CVD risk among adults with comorbid HIV and hypertension. The intervention was developed by adapting an evidence-based intervention using the ADAPT-ITT framework and integrating formative community-based data collection. The adaptation evolved significantly through iterative input from an intervention design team of diverse stakeholders from Tanzania. A pilot feasibility trial of the intervention was initiated in December 2024 (ClinicalTrials.gov, NCT06503991) [35]. The trial will produce further evidence on the feasibility and acceptability of the *COACH* intervention, as well as its potential impact on clinical outcomes of hypertension control and care engagement.

## Declarations

*Ethics approval and consent to participate*: The study was performed in line with the principles of the declaration of Helsinki and received ethical approval from institutional review boards at Duke Health (Pro00090902), KCMC (No. 1454), and the Tanzania National Institute of Medical Research (NIMR/HQ/R.8a/Vol.IX/4615).

## Consent for publication

Not applicable.

## Availability of data and materials

Data is provided within the manuscript or supplementary information files. The data are available from the authors upon reasonable request.

## Authors contributions

PM drafted this manuscript. PM, JTH, BTM and NMT implemented and supervised the study. PM, JTH, BTM, NTM, THJ, HBB, and FMS contributed to development of the study concept and design. LW, KFH, GK, JM, PS, and SFS contributed to administration and coordination of the study and data collection. PM, JTH, and NMT analyzed the data. PM, BTM, NMT, MHW, SFS, THJ, HBB, FMS, LW, KFH, GK, JM, PS and JTH all contributed to the interpretation of the data, made critical revisions to the article and read and approved the final article draft.

## Data Availability

All data produced in the present work are contained in the manuscript

## Acknowledgements

We would like to acknowledge support received by the Duke Global Health Institute, the Kilimanjaro Clinical Research Institute and the KCMC-Duke Collaboration. We gratefully acknowledge the contributions of Dr. Anzibert Rugakingira, Dr. Remyda Zebedayo, Dr. Francis Gwasma and the staff and patients of the study sites, Majengo Care and Treatment Center and Pasua Care and Treatment Center, who contributed to the development of the intervention.

