## Supplementary Material for "Adapting an intervention to improve hypertension care for adults with HIV in Tanzania: Co-design of the Community Health Worker Optimization of Antihypertensive Care in HIV (COACH) intervention"

**CHW COUNSELING CLINIC SESSION 1 – EDUCATION SESSION (20 minutes)**

**INTRODUCTION**

- Introduce yourself to the patient
- ***You are here with us participating in the intervention because you have high BP.***
- *The goal of the education sessions is to empower you by increasing your understanding of HTN, promoting self-management practices, and fostering healthy behaviors.*

**BLOOD PRESSURE VALUE DISCUSSION**

- The RA will record the patient’s BP value on the patient handout and then give the patient handout to the CHW for the session.
- The CHW will record the patient’s BP value in the tracking log
- Tell the patient their value today.
- Note that you will discuss more about what the numbers mean shortly.

*Please feel free to ask questions any time.*

**DEFINITION OF HIGH BLOOD PRESSURE**

- *High BP is when the pressure of blood moving through the body is high* – **No further explanation (use visual aid #1).**
- High blood pressure is the same as hypertension (shinikizo la juu la damu).
- High BP is a **chronic disease** – **a disease you will have the rest of your life** – like diabetes or HIV. It is not temporary.
- High BP is not thinking too much. It is not receiving shocking news or being angry or being stressed.
- High BP is not when the heart is beating too fast.
- Patients rarely feel symptoms from high blood pressure.

**HEALTHY BLOOD PRESSURE LEVELS**

- ***Everyone has a BP (i.e. pressure of blood moving through the body). Having a BP is normal. Some people have a normal BP, but some other people may have a high BP***
- Use visual aid #2 –yako to discuss normal (systolic 139 or lower *AND* diastolic 89 or lower) and hypertensive (systolic 140 or higher *OR* diastolic 90 or higher)

**THE CAUSES OF HIGH BP**

- **Major causes of high BP include aging, family history, being overweight, being physically inactive, and eating a diet high in salts, fats/oils and sugars, smoking and drinking alcohol (use visual aid #3-8).**
- **Some people may not have any of these risk factors and still have hypertension. This is common and normal, because hypertension is a very common condition.**
- *Many people think that thinking too much, worrying, being angry & receiving shocking news are* ***THEIR*** *causes of high BP but this is not true. While stress is not good for our body, it is not a* ***MAJOR*** *cause of high BP.*
- *HIV and HIV medications do not cause hypertension.*

**WHAT ARE THE WARNING SIGNS OF HIGH BP?**

- *Most people with high BP have no symptoms unless it is very severe.*
- *They feel completely well.*
- *The only way to know if your BP is high is to check it instead of relying on symptoms to alert you.*

**WHAT ARE THE RISKS OF HAVING UNTREATED HIGH BLOOD PRESSURE?**

- **Having untreated high BP for a long time will lead to complications. These complications include stroke, heart attack, heart failure, kidney failure, vision problems, and erectile dysfunction - use visual aid #9**
- *Some people think that these complications are the signs and symptoms of high BP but they are not. They are complications that occur from having untreated high BP for a long time.*
- As BP increases, the risk of complications also increase
- ***People cannot fall down suddenly or die suddenly from high BP. Someone with high BP can die only after having untreated high BP for a long time.***
- ***Controlling high blood pressure is a very important step to reduce risk of these complications. This is why you should take medications.***

**INTRODUCE THE IDEA OF TREATMENT**

- *High BP can be controlled by using medication every day for the rest of your life & lifestyle changes just as it is with your HIV care.*
- *This is the only way to control your high BP* – use visual aid #10 to describe similarities between HTN and HIV care
- Advise the patient to see their doctor monthly to monitor and treat their high BP.
- **You may be prescribed multiple medications to control your BP. This is very common.**
- *The dose of the medication may increase and this is also very common.*
- Medications are the best and fastest way to control high BP. Lifestyle changes take a long time to work but are also very important for your health.
- If prescribed medications, they must be used every day for the rest of your life. ***The correct BP medications will not make your BP too low; they will control your BP so it stays at a normal range.***
- **You cannot cure hypertension.** Hypertension requires lifelong treatment with BP medications.
  - Stopping thinking too much does not cure hypertension
  - Going home and resting does not cure hypertension
  - Using herbal treatment does not cure hypertension
  - Garlic, black seed, and drinking lots of water do not treat hypertension
  - Using medications for only a short amount of time does not cure hypertension

**COMMIT TO STARTING TREATMENT**

- **There is a very high chance that you will be prescribed BP medications today (if you are not already on medication).**
- ***I will refer you to the doctor in a few minutes. It's important to see the doctor today so they can review your progress and make any necessary adjustments to your medications or provide new prescriptions. This follow-up is vital for managing your health effectively.***
- *Next week, I will call you to hear from you how the consultation went and discuss more on the medicines that you will be prescribed. We will meet again in the clinic next month*.
- *Record the patient’s phone number and clinic appointment dates on the patient handout and tracking log*.
- *Remind the patient to bring their patient handout in the next clinic appointment.*
- Ask the patient what questions they have.

**CHW COUNSELING CLINIC SESSION 2 – HYPERTENSION TREATMENT (30 minutes)**

**INTRODUCTION**

- Greet the patient
- What questions do you have before beginning the session?

**SESSION 1 BRIEF REVIEW –** *Before we begin the session today let’s review a few things from our last session.*

- **High BP is defined by what values?** – *A high BP value is any value 140 or greater OR any value 90 or greater. A normal BP is any systolic value 139 or lower AND any diastolic value 89 or lower. Use visual aid #2*
- **What causes high BP?** – use visual aids #3-8 to briefly discuss 1) aging, 2) family history, 3) obesity, 4) sedentary lifestyle, 5) salty and unhealthy diet, 6) smoking and alcohol are the major causes of high BP.
- **What does not cause high BP?** Overthinking, anger, stress, and receiving shocking news are not **YOUR** causes of high BP.
- **What are the signs and symptoms of High BP?** – *High BP has no signs or symptoms unless severe. Most people with high BP feel completely well. The only way to know if your BP is high is to check it*
- **What are the complications of having untreated high BP for a long time?** – *Having untreated high BP for a long time can lead to complications*. Use visual aid #9
- **How can you treat high BP?** – *BP medications taken every day for life and lifestyle modifications are the only way to control high BP.*
- If the patient drank coffee, did an exercise or smoked, ask to see whether they have a 30 minutes gap between the activity and the time you want to measure their blood pressure.
- If SBP ≥ 180 **OR** DBP ≥ 120 **and** the patient is experiencing symptoms such as severe headache, confusion, chest pain or difficulty in breathing; refer them to the doctor immediately. Do not proceed with the counselling session.
- **If the patient is pregnant, notify the research assistant and the provider. The patient should not continue in the study.**

**BLOOD PRESSURE VALUE DISCUSSION**

- Record the patient’s BP value on the patient handout and tracking log.
- State what the blood pressure is this visit, and how this compares to the blood pressure from the last visit
- Comment on whether the patient’s blood pressure value meets the goal blood pressure, **LESS THAN 140/90. Congratulate patient if their value is below 140/90.**

**SCENARIO 1: NORMAL BP (SBP ≤ 139 and DBP ≤ 89) AND NOT PRESCRIBED BP MEDICATION**

**LIFESTYLE MODIFICATION COUNSELING – REFER TO PAGE 7 (CLINIC SESSION 3)**

- **Stress importance of BP monitoring and likelihood of needing medication in the future.** BP fluctuates all the time and increases with age so there’s a high chance your BP will be high in the future.
- Using medications to control BP is very common & if prescribed medications you will need to use them for the rest of your life just like with HIV. **The correct** **BP medications will not make your BP too low; it will control your BP so it stays at a normal range.**

**SCENARIO 2: NORMAL BP (SBP ≤ 139 and DBP ≤ 89) AND PRESCRIBED BP MEDICATIONS**

**ASSESS PRIOR TREATMENT**

- *Did the doctor prescribe BP medications?*
- *How many times in a week do you typically miss your dose?*
- *Can you tell me all the reasons for missing the dose?*
- **Brainstorm with the patient for solutions to improve BP medication adherence.** **Here are some tips to improve your drug adherence;**
  - Set alarms on your phone to alert you when it is time to take your medication. Help the patient set a daily alarm on their phone if they are interested.
  - Try to take your medications at the same time each day perhaps linked into your daily routine. For example, taking it when the news starts at 8.00 pm, right after brushing your teeth, right after a meal, or when taking your ART.
  - It is okay and safe to take your blood pressure medications the same time you take your HIV medications.
  - Store your BP medications with your ART so that you see all your medications together and remember to take them both.
- *Remember, taking your medication as prescribed is one of the most important things you can do to manage your health and prevent complications. You are doing a great job by taking steps to improve your adherence.*
- If you experience any side effects while using BP medications, do not stop it. Instead, talk to your doctor on ways to mitigate or manage these side effects.
- **Coming to the clinic for your drug refills and monthly blood pressure monitoring is crucial to ensure your treatment is working effectively and to monitor your health closely.**
- Maintaining a healthy lifestyle through diet, regular exercise, and stopping smoking and alcohol is also crucial to keeping your blood pressure within a normal range. We will spend more time discussing this in our next session.
- ***Remember that decreasing your thoughts, stopping thinking, drinking lots of water and lying down and resting are not ways to control high BP. This is because thinking too much and worrying is not a cause of high BP. The only way to control high BP is through lifestyle changes and BP medications.***
- ***You cannot cure high BP by using medications for a short time.***
- ***You cannot cure high BP by chewing herbs or spices such as garlic or black seed.***
- ***Remember you can take your medication at any time of day or night (so it will not conflict with fasting).***
- ***Your blood pressure medications can safely be taken either on an empty stomach or with any food or drink (including milk).***
- ***I will refer you to the doctor in a few minutes. It's important to see the doctor today so they can review your progress and provide new prescriptions. This follow-up is vital for managing your health effectively.***
- *Record the patient’s phone and clinic appointment dates on the patient handout and tracking document*.
- *Remind the patient to bring their patient handout in the next clinic appointment.*
- Ask the patient what questions they have before the session ends.

**SCENARIO 3: HIGH BP (SBP ≥ 140 and DBP ≥ 90) AND NOT PRESCRIBED BP MEDICATIONS**

**ASSESS PRIOR TREATMENT**

- *What are you currently doing to control your high blood pressure?*
- *Can you share any thoughts or feelings you have about using medications to control your high BP?*
- **There is a very high chance that you will be prescribed BP medications today, here are some tips to improve your drug adherence:**
  - Set alarms on your phone to alert you when it is time to take your medication
  - Try to take your medications at the same time each day perhaps linked into your daily routine. For example, taking it when the news starts at 8.00 pm, right after brushing your teeth, or right after a meal, or when taking your ART.
  - It is okay and safe to take your blood pressure medications the same time you take your HIV medications.
  - Store your BP medications with your ART so that you see all your medications together and remember to take them both.
- If you experience any side effects while using BP medications, don’t stop it. Instead, talk to your doctor on ways to mitigate or manage these side effects.
- **Using medications to control BP is very common & if prescribed medications you will need to use them for the rest of your life just like with HIV. The correct BP medications will not make your BP too low; it will control your BP so it stays at a normal range.**
- ***Remember that decreasing your thoughts, stopping thinking, drinking lots of water and lying down and resting are not ways to control high BP. This is because thinking too much and worrying is not a cause of high BP. The only way to control high BP is through lifestyle changes and by taking BP medications every day.***
- ***You can’t cure high BP by using medications for a short time.***
- ***You can’t cure high BP by chewing herbs or spices such as garlic or black seed***
- ***I will refer you to the doctor in a few minutes. It's important to see the doctor today so they can review your progress and provide new prescriptions. This follow-up is vital for managing your health effectively.***
- *Next week, I will call you to hear from you how the consultation went and discuss more on the medicines that you will be prescribed. We will meet again in the clinic next month*.
- *Record the phone and clinic appointment dates on the patient handout and tracking log*.
- *In the next session, we will go over lifestyle changes that will help to control your BP and review medications.*
- *Remind the patient to bring their patient handout in the next clinic appointment.*
- Ask the patient what questions they have before the session ends.

**SCENARIO 4: HIGH BP (SBP ≥ 140 and DBP ≥ 90) AND PRESCRIBED BP MEDICATIONS**

**ASSESS PRIOR TREATMENT**

- *Did the doctor prescribe BP medications?*
- *How many times in a week do you typically miss your dose?*
- *Can you tell me all the reasons for missing the dose?*
- **It is normal for patients with high blood pressure medications to need their medications and/or doses changed to improve their blood pressure. Do not be discouraged that your BP is elevated today despite taking medications.**
- **If taking BP medications daily as prescribed: There is a very high chance that you will be prescribed different BP medications today or the dose of your BP medication might be increased, this is normal.**
- **If not taking BP medications daily as prescribed: Here are some tips to help to remember to take your BP medications everyday:**
  - Set alarms on your phone to alert you when it is time to take your medication. Help the patient set a daily alarm on their phone if they are interested.
  - Try to take your medications at the same time each day perhaps linked into your daily routine. For example, taking it when the news starts at 8.00 pm, right after brushing your teeth, or right after a meal, or when taking your ART.
  - It is okay and safe to take your blood pressure medications the same time you take your HIV medications.
  - Store your BP medications with your ART so that you see all your medications together and remember to take them both.
  - Ask a family member or friend to remind you or check in with you to see if you’ve taken your medications
- If you experience any side effects while using BP medications, don’t stop it. Instead, talk to your doctor on ways to **mitigate** or manage these side effects.
- **Using medications to control BP is very common & if prescribed medications you will need to use them for the rest of your life just like with HIV. The correct BP medications will not make your BP too low; it will control your BP so it stays at a normal range. Coming to the clinic for your drug refills and monthly blood pressure monitoring is crucial to ensure your treatment is working effectively and to monitor your health closely.**
- ***Remember that decreasing your thoughts, stopping thinking, drinking lots of water and lying down and resting are not ways to control high BP. This is because thinking too much and worrying is not a cause of high BP. The only way to control high BP is through lifestyle changes and BP medications.***
- ***You can’t cure high BP by using medications for a short time.***
- ***You can’t cure high BP by chewing herbs or spices such as garlic or black seed***
- ***I will refer you to the doctor in a few minutes. It's important to see the doctor today so they can review your progress and make any necessary adjustments to your medications or provide new prescriptions. This follow-up is vital for managing your health effectively.***
- *Next week, I will call you to hear from you how the consultation went and discuss more on the medicines that you will be prescribed. We will meet again in the clinic next month*.
- *Record the phone and clinic appointment dates on the patient handout and tracking log.*
- *Remind the patient to bring their patient handout in the next clinic appointment.*
- Ask the patient what questions they have before the session ends.

**CHW COUNSELING CLINIC SESSION 3 – LIFESTYLE CHANGES (30 minutes)**

**INTRODUCTION**

- Greet the patient
- Ask what questions the patient has before beginning the session

**IMPORTANT INSTRUCTIONS FOR BLOOD PRESSURE MEASUREMENTS.**

- See instructions on the poster on the wall or refer back to clinic session 2 on page 3 for complete instructions.

**BLOOD PRESSURE VALUE DISCUSSION**

- State what the blood pressure is this visit, and how this compares to previous visits and record the value on the patient handout and in the tracking document
- Comment on whether or not the patient is at goal – **BP LESS THAN 140 AND LESS THAN 90. Congratulate the patient if their BP is less than 140/90.**
- Provide encouragement – Lifestyle changes improve BP slowly over time. Medications improve BP faster but only *if* you are on the correct medication and dose

**ASSESS PATIENT ADHERENCE TO BP MEDICATION**

- *Did the doctor prescribe BP medications?*
- *How many times in a week do you typically miss your dose?*
- *Can you tell me all the reasons for missing the dose?*
- ***Your blood pressure medications are not addictive.***

**SESSION 2 BRIEF OVERVIEW**

- *Using blood pressure medications, using multiple blood pressure medications, and increasing the dose of blood pressure medication over time is normal & common*
- *You will need to use the BP medication every day for the rest of your life just like your HIV medication – use visual aid #10*
- *It is important to check your BP and visit the doctor monthly to make sure that your BP is controlled and that you are on the right medications and at the right dose.* ***When used and monitored correctly, these*** ***medications will not make your BP too low; they will control your BP so that it stays at a normal range.***
- ***Remember that stopping thinking and resting is not a way to control high BP. This is because thinking too much, stress, receiving shocking news and anger are not a cause of hypertension. High BP cannot be cured.***
- ***Eating natural herbs and spices like garlic and black seed cannot cure High BP.***
- ***You cannot cure High BP by using medications for a short period of time.***
- ***You should not stop your BP medications once your blood pressure has normalized.***
- ***Hypertension is a chronic illness. The only way to control hypertension is through BP medications and lifestyle changes.***

**LIFESTYLE MODIFICATION COUNSELING**

- *Lifestyle changes can also help to reduce blood pressure*
- Lifestyle changes must be used with medications **and cannot replace medications**

1. SALT:

- **Reducing salt will improve your BP. Use visual aid #5**
- *Here are some ways to reduce salt intake:*
- *Cook with less salt –* ***don’t use more than a pinch of salt when cooking***
- *Don’t add salt to food that is already prepared. Don’t have a salt shaker on the table*
- *Eat healthier meats like chicken and fish. Reduce intake of beef, pork and goat*
- *Limit the consumption of salty snacks including chips, crisps and crackers*
- *Avoid most restaurant food especially chips, nyama choma, fried ndizi, chapati, cakes, mandazi, and other fried foods. See visual aid for foods to avoid.*
- *Flavor your food with garlic, ginger, lemon juice, lime, onion, vinegar, herbs and spices like pepper, chili, turmeric, jeera, curry powder, cardamom, and cloves. Avoid chili salt, ketchup, and beef and chicken cubes*

2. WEIGHT LOSS:

- **If you are at an unhealthy weight, then weight loss can help to reduce blood pressure. Use visual aid #13 –**
- *Do you think that you are at a healthy or unhealthy weight?*
- Maintaining a healthy weight will help control BP. But even losing a few kilos will help to reduce BP
- **Best way to lose weight OR to maintain a healthy weight is to eat the correct portions of food, eat a healthy diet, and exercise**
- Show patient a **plate** and discuss portion control using **visual aids #12**
- **Reduce**:
  - *Rice*
  - *Ugali*
  - *Chapati*
  - *Bread*
- **Stop**:
  - *Stop drinking juice, soda, malt beverages (filled with sugar), alcohol* ***(makes you gain a lot of weight and causes high BP)***
  - *Stop using sugar – sugary foods make you gain weight and are unhealthy*
  - *Fried and oily foods*
  - *Juice is not a fruit and will make you gain weight*
- **Increase***:*
  - *Water (at least 2 liters per day)*
  - *Healthy fruits and vegetables – eating healthy fruits and vegetables will help control your BP*

3. EXERCISE:

- **Exercise will help to control your BP. Use visual aid #14**
- *Tell me about the exercise or movement you get in a typical day.*
- Engage in activity at least 30 minutes most days of the week
- Activities should increase your heart rate, make you sweat and make you breathe hard

4. SMOKING CESSATION:

- **Smoking cigarettes & using tobacco causes high BP & damage to your body. Use visual aid #8** If smokes/uses tobacco, advise to try and completely stop

5. ALCOHOL MODERATION:

- **Excess use of alcohol can also cause high blood pressure. Use visual aid #15**
- *Do you drink alcohol?*
- *How many days a week do you drink?*
- *How many drinks per day?*
- *If you drink alcohol, you should try and stop or reduce alcohol because drinking alcohol causes high BP*

**PLAN OF ACTION**

- *What are your thoughts about what we just discussed?*
- *Are there any lifestyle changes that you would like to make from these choices?*

Choose from:

- Weight loss or maintaining a healthy weight - eating a healthy diet with appropriate portions
- Salt reduction
- Exercise
- Stopping smoking / using tobacco
- Stopping alcohol

**Do not force the patient to choose any particular change.**

For each change:

- *How important is it to you to make this change?*
- *How confident are you that you can make that change?* **Help patient come up with a specific and concrete plan of action**
- *What do you think might be difficult about making that change?*
- Record the lifestyle changes the patient has committed to and their plan on ‘**Handout**’

**CLOSING AND GOAL SETTING**

- Encourage monthly follow-up and blood pressure checks with the doctor.
- Encourage honesty and openness with the doctor.
- *Lifestyle changes help to control blood pressure but usually take a while to work (many months). Sometimes lifestyle changes are not enough to control blood pressure and people will also need medications.* ***If you are prescribed medications, lifestyle changes cannot replace the medications to control your BP****. If you are prescribed medications, you will need to use them for the rest of your life. This is okay and is very normal*
- Ask what questions they have and if they would like any additional information before referring them to the doctor.
- *Record the patient’s phone and clinic appointment dates on the patient handout and tracking log*
- *Remind the patient to bring their patient handout in the next clinic appointment.*

**CHW COUNSELING CLINIC SESSION 4 – HYPERTENSION EDUCATION (20 minutes)**

**INTRODUCTION**

- Greet the patient
- Ask what questions the patient has before beginning the session

**IMPORTANT INSTRUCTIONS FOR BLOOD PRESSURE MEASUREMENTS.**

- See instructions on the poster on the wall or in clinic session 2 on page 3

**BLOOD PRESSURE VALUE DISCUSSION**

- State what the blood pressure is this visit, and how this compares to previous visits. Record the blood pressure on the patient handout and in the tracking document
- Comment on whether or not the patient is at goal – **BP LESS THAN 140 AND LESS THAN 90. Congratulate patient if value is normal.**
- Provide encouragement – Lifestyle changes improve BP slowly (months). Medications improve BP faster but only *if* you are on the correct medication and dose

**ASSESS PATIENT ADHERENCE TO BP MEDICATION**

- *Did the doctor prescribe BP medications?*
- *How many times in a week do you typically miss your dose?*
- *Can you tell me all the reasons for missing the dose?*
- *Have there been any changes in your health or new symptoms since we last spoke?*
- *Using blood pressure medications, using multiple blood pressure medications, and increasing the dose of blood pressure medication over time is normal & common*
- *You will need to use the BP medication every day for the rest of your life just like your HIV medication – use visual aid #10*
- *It is important to check your BP and visit the doctor monthly to make sure that your BP is controlled and that you are on the right medications and at the right dose.* ***When used and monitored correctly, these*** ***medications will not make your BP too low; they will control your BP so that it stays at a normal range.***
- ***Remember that stopping thinking and resting is not a way to control high BP. This is because thinking too much, stress, receiving shocking news and anger are not causes of hypertension. High BP cannot be cured.***
- ***Eating natural herbs and spices like garlic and black seed cannot cure High BP.***
- ***You cannot cure High BP by using medications for a short period of time.***
- ***You should not stop your BP medications once your blood pressure has normalized.***
- ***Hypertension is a chronic illness. The only way to control hypertension is through BP medications and lifestyle changes.***

**LIFESTYLE MODIFICATION CHANGES FOLLOW UP**

- How is it going with the changes that you chose to work on in the past month?
- What challenges have you experienced while making those changes?
- Encourage and work with the patient to come up with solutions to the challenges they are experiencing.
- Are there any new changes that you would like to work on?

**HEALTH EDUCATION – *Today we will be discussing hypertension causes and warning signs in more detail.***

**THE CAUSES OF HIGH BP**

- Aging – *The elderly are at increased risk of high BP. As you age your risk of high BP increases.* **DO NOT MENTION SPECIFIC AGES**
- Family history
- Being overweight – use visual aid #3
- Being physically inactive – use visual aid #4
- Eating a diet high in ***salt***, fats, sugar – use visual aid #5-6
- Smoking – use visual aid #7
- Drinking alcohol – use visual aid #8
- Some people with hypertension may have none of these risk factors for hypertension. This is very normal and common because hypertension is a very common disease.
- *Many people think that thinking too much, worrying, being angry & receiving shocking news are* ***THEIR*** *causes of high BP but this is not true. While stress is not good for our body it is not a* ***MAJOR*** *cause of high BP. What are your thoughts about that?*
- *When people are diagnosed with HIV and learn about their status, they often think that receiving this shocking news can cause high blood pressure, but this is not true. They actually may have had high BP for a long time, before they were diagnosed with HIV. But they were only told they had high BP after they were diagnosed with HIV because this is when their BP was checked for the first time.*
- *HIV and HIV medications do not cause high BP.*
- What questions do you have?

**WHAT ARE THE WARNING SIGNS AND COMPLICATIONS OF HIGH BP?**

- *Most people with high BP have no symptoms unless it is very severe. They feel completely well.*
- *The only way to know if your BP is high is to check it instead of relying on symptoms to alert you.*
- *Common complications of long-term uncontrolled hypertension are strokes, blindness, heart attacks, kidney failure. Use visual aid #9*

**KEY REMINDERS AND CLOSING**

- Hypertension is a chronic condition that needs ongoing management, similar to HIV.
- Having untreated high BP for a long time leads to complications. Managing high BP helps prevent these serious complications and maintain overall health. Use visual aid #9
- Ask the patient if they have any questions before referring them to the clinician.
- *Record the patient’s phone and clinic appointment dates on the patient handout and tracking log.*
- *Remind the patient to bring their patient handout in the next clinic appointment.*

**CHW COUNSELING CLINIC SESSION 5 – MEDICATION & LIFESTYLE CHANGES FOLLOW UP (20 minutes)**

**INTRODUCTION**

- Greet the patient
- Ask what questions the patient has before beginning the session

**IMPORTANT INSTRUCTIONS FOR BLOOD PRESSURE MEASUREMENTS.**

- See instructions on the poster on the wall or in clinic session 2 on page 3.

**BLOOD PRESSURE VALUE DISCUSSION**

- State what the blood pressure is this visit, and how this compares to previous visits. Record the blood pressure on the patient handout and tracking document
- Comment on whether or not the patient is at goal – **BP LESS THAN 140 AND LESS THAN 90**

Provide encouragement – Lifestyle changes improve BP slowly (months). Medications improve BP faster but only *if* you are on the correct medication and dose

**ASSESS PATIENT ADHERENCE TO BP MEDICATION AND LIFESTYLE CHANGES**

***Adhering to BP medications has many benefits including*** ***maintaining a healthy blood pressure level and reducing the risk of complications***.

- *Did the doctor prescribe BP medications?*
- *How many times in a week do you typically miss your dose?*
- *Can you tell me all the reasons for missing the dose?*
- **Brainstorm with the patient for solutions to improve BP medication adherence. Here are some tips to improve your drug adherence;**
  - Set alarms on your phone to alert you when it is time to take your medication. Help the patient set a daily alarm on their phone if they are interested.
  - Try to take your medications at the same time each day perhaps linked into your daily routine. For example, taking it when the news starts at 8.00 pm, right after brushing your teeth, right after a meal, or when taking your ART.
  - It is okay and safe to take your blood pressure medications the same time you take your HIV medications.
  - Store your BP medications with your ART so that you see all your medications together and remember to take them both.
- *Remember, taking your medication as prescribed is one of the most important things you can do to manage your health and prevent complications. You are doing a great job by taking steps to improve your adherence.*
- If you experience any side effects while using BP medications, do not stop it. Instead, talk to your doctor on ways to mitigate or manage these side effects.
- **Coming to the clinic for your drug refills and monthly blood pressure monitoring is crucial to ensure your treatment is working effectively and to monitor your health closely.**
- Maintaining a healthy lifestyle through diet, regular exercise, and stopping smoking and alcohol use is also crucial to keeping your blood pressure within a normal range.
- ***Remember that decreasing your thoughts, stopping thinking, drinking lots of water and lying down and resting are not ways to control high BP. This is because thinking too much and worrying is not a cause of high BP. The only way to control high BP is through lifestyle changes and BP medications.***
- ***You cannot cure high BP by using medications for a short time.***
- ***You cannot cure high BP by chewing herbs or spices such as garlic or black seed.***
- ***Remember you can take your medication at any time of day or night (so it will not conflict with fasting).***
- ***Your blood pressure medications can safely be taken either on an empty stomach or with any food or drink (including milk).***
- Ask the patient if they have any questions related to BP medications.
- Can you share your experience with me in trying to limit salt, oils/fats or sugar in your diet?
- Encourage and work with the patient to come up with solutions to the challenges they are experiencing.
- What is your current exercise routine? Provide encouragement if they exercise about 30 minutes per day. If not, work with the patient to see how they best they can engage in physical activity. Suggest participating in activities such as brisk walking, jogging, dancing (e.g. rumba), cycling, local sports such as football & netball.
- **If they smoke or use chewing tobacco:**
  - How is it going with reducing/quitting smoking or tobacco?
  - Can you tell me how much cigarettes you have smoked or tobacco you have used this past week?
  - What challenges are you experiencing with stopping smoking or tobacco?
- **If they drink:**
  - How is it going with cutting down on alcohol use?
  - Can you tell me how much alcohol you have been taking this past week?
  - Are there any challenges with reducing your alcohol intake?
- *Encourage and work with the patient to come up with solutions to the challenges they are experiencing.*

**SESSION 4 BRIEF REVIEW –** *Let’s review a few things from our last session. Ask the patient to remind you of the topics below. Remind them if they have forgotten and address ALL misconceptions raised by the patient.*

- **Which values suggest high BP?** – *A high BP value is any value 140 or greater OR any value 90 or greater. A normal BP is any systolic value 139 or lower AND any diastolic value 89 or lower. Use visual aid #***What are the Signs and symptoms of High BP?** – *High BP has no signs or symptoms unless severe. Most people with high BP feel completely well. The only way to know if your BP is high is to check it*
- **What are the Complications of having untreated High BP for a long time?** – *Having untreated high BP for a long time can lead to complications*. Use visual aid #9**How can you treat High BP?** – *Lifestyle changes and BP medications taken every day for life are the only way to control high BP. We will spend some time discussing medications now*

**ESTABLISHING LONG-TERM TREATMENT GOALS**

- Are there any areas where you would like more information or support?
- In the next 2 months, the study will be over and you will no longer receive free BP medications, and you will need to be buying them. What strategies do you have in mind for covering the cost of your BP medication moving forward?
- Let’s talk about any potential challenges you might face in the future and come up with strategies to address them.
- It is crucial to continue taking your medications everyday as prescribed by your provider and sticking to the lifestyle changes that you have chosen, as well as monitoring your BP monthly to help you keep your blood pressure under control and reduce your risk of developing complications.

**KEY REMINDERS & CLOSING**

- Ask patients if they have any questions or need clarification on anything related to hypertension before referring them to the doctor.
- *Record the patient’s phone and clinic appointment dates on the patient handout and tracking log*.
- *Remind the patient to bring their patient handout in the next clinic appointment.*

**CHW COUNSELING CLINIC SESSION 6 – FINAL SESSION (20 minutes)**

**INTRODUCTION**

- Greet the patient
- Ask what questions the patient has before beginning the session

**BLOOD PRESSURE VALUE DISCUSSION**

- The RA will record the patient’s BP value on the patient handout and then give the patient handout to the CHW for the session.
- State what the blood pressure is this visit, and how this compares to previous visits and record value in tracking log.
- Comment on whether or not the patient is at goal – **BP LESS THAN 140 AND LESS THAN 90. Congratulate the patient if their BP is controlled**
- Provide encouragement – Lifestyle changes improve BP slowly (months). Medications improve BP faster but only *if* you are on the correct medication and dose
- ***Monitoring your BP consistently is important to ensure that your BP maintains the goal.***

**ASSESS PATIENT ADHERENCE TO BP MEDICATION AND LIFESTYLE CHANGES**

***Adhering to BP medications has many benefits including maintaining a healthy BP level and reducing the risk of complications.***

- *Did the doctor prescribe BP medications?*
- *How many times in a week do you typically miss your dose?*
- *Can you tell me all the reasons for missing the dose?*
- *Have there been any changes in your health or new symptoms since we last spoke?*
- **Brainstorm with the patient for solutions to improve BP medication adherence. Here are some tips to improve your drug adherence;**
  - Set alarms on your phone to alert you when it is time to take your medication.
  - Try to take your medications at the same time each day perhaps linked into your daily routine. For example, taking it when the news starts at 8.00 pm, right after brushing your teeth, right after a meal, or when taking your ART.
  - It is okay and safe to take your blood pressure medications the same time you take your HIV medications.
  - Store your BP medications with your ART so that you see all your medications together and remember to take them both.
- *Remember, taking your medication as prescribed is one of the most important things you can do to manage your health and prevent complications. You are doing a great job by taking steps to improve your adherence.*
- If you experience any side effects while using BP medications, do not stop it. Instead, talk to your doctor on ways to mitigate or manage these side effects.
- **Coming to the clinic for your drug refills and monthly blood pressure monitoring is crucial to ensure your treatment is working effectively and to monitor your health closely.**
- Maintaining a healthy lifestyle through diet, regular exercise, and stopping smoking and alcohol use is also crucial to keeping your blood pressure within a normal range.
- ***Remember that decreasing your thoughts, stopping thinking, drinking lots of water and lying down and resting are not ways to control high BP. This is because thinking too much and worrying is not a cause of high BP. The only way to control high BP is through lifestyle changes and BP medications.***
- ***You cannot cure high BP by using medications for a short time.***
- ***You cannot cure high BP by chewing herbs or spices such as garlic or black seed.***
- ***Remember you can take your medication at any time of day or night (so it will not conflict with fasting).***
- ***Your blood pressure medications can safely be taken either on an empty stomach or with any food or drink (including milk).***
- Ask the patient if they have any questions related to BP medications
- Assess the patient’s progress with lifestyle changes. Review any challenges they’ve encountered and celebrate any successes.

**ESTABLISHING LONG-TERM TREATMENT GOALS**

- *As this is our final session, I want to make sure you're feeling confident and prepared to manage your hypertension on your own. Last time, we talked about your long-term treatment goals, including understanding your condition, staying on top of your medication and lifestyle changes, and understanding the complications of uncontrolled hypertension. Today, let's review everything in detail.*
- **Which values suggest high BP?** – *A high BP value is any value 140 or greater OR any value 90 or greater. A normal BP is any systolic value 139 or lower AND any diastolic value 89 or lower. Use visual aid #***What are the Signs and symptoms of High BP?** – *High BP has no signs or symptoms unless severe. Most people with high BP feel completely well. The only way to know if your BP is high is to check it.*
- **What are the Complications of having untreated High BP for a long time?** – *Having untreated high BP for a long time can lead to complications*. Use visual aid #9
- **How can you treat High BP?** – *Lifestyle changes and BP medications taken every day for life are the only way to control high BP. We will spend some time discussing medications now.*
- Are there any areas where you would like more information or support?
- Have you thought of any strategies for paying for your BP medications moving forward as we discussed last month?
- Let’s talk about any potential challenges you might face in the future and come up with strategies to address them.
- It is crucial to continue taking your medications everyday as prescribed by your provider and sticking to the lifestyle changes, as well as monitoring your BP and seeing your doctor monthly to keep your blood pressure under control and reduce your risk of developing complications.
- Ask if the patient has any questions or concerns. *I’m here to help with any last-minute support or clarifications that you need.* **Address all misconceptions that the patient has about hypertension or its management including garlic, black seed, water, thinking too much, milk, erectile dysfunction, etc.**
- Encourage routine follow-up with the doctor, checking BP routinely, and encouraging honesty and openness with the doctor.

**CLOSING REMARKS.**

- Ask the patient if they have any questions.
- ***You’ve done a great job participating in these sessions. Keep up the good work, and don’t hesitate to reach out if you need any further support.***
- Refer patient to the doctor.

**CHW TELEPHONE SESSIONS 1 – 4**

**INTRODUCTION**

- Introduce yourself to the patient
- Ensure this is a convenient time to talk with the patient

**QUESTIONS**

- *Do you have any questions about your blood pressure or your medications since our last conversation?*
- *How are things going with your blood pressure medication?*
- *Have you experienced any challenges or issues with taking it?*
  - **If there are challenges,** discuss potential solutions*: I understand. Let us think about some potential solutions that might help. What do you think could make it easier for you to take your medication as prescribed?*
  - **If there are no challenges,** provide positive reinforcement: *That’s great to hear. Keep up the good work!*
- Briefly discuss benefits of medication compliance

**APPOINTMENT REMINDER**

- Remind patient the only way to treat high BP is with medications and lifestyle changes. Thinking too much does not cause high BP. High BP cannot be cured.
- Medications are the best and fastest way to control BP.
- Ask what questions the patient has – correct **ALL misconceptions** that the patient raises.
- *Just a quick reminder that your next appointment is on* ***XXXX.*** *We look forward to seeing you them. Please bring your handout along.*
- *Thank you for your time and don’t hesitate to call me if you have any questions or concerns before our next meeting. Have a great day!*

**CHW CLINIC SESSIONS 1 – 6: ACCOMPANYING VISUAL AID IMAGES FOR COUNSELING SESSIONS**

**1.**

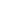

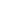

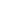

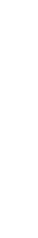

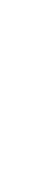

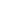

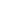

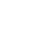

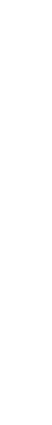

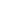

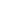

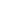

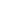

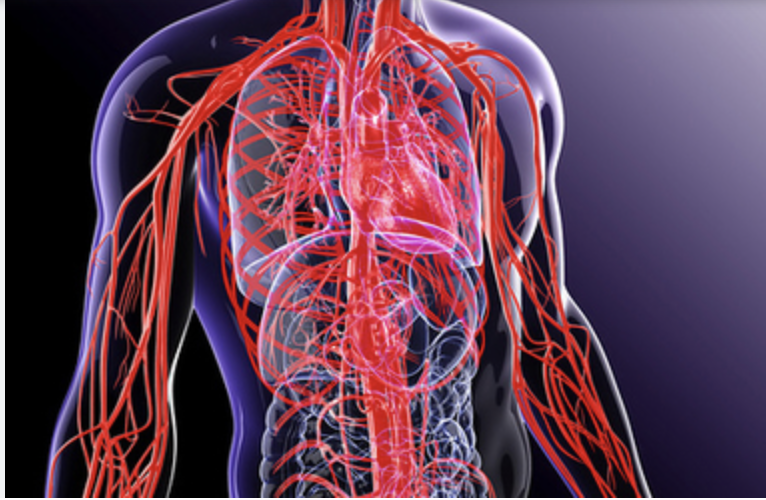

**3.**

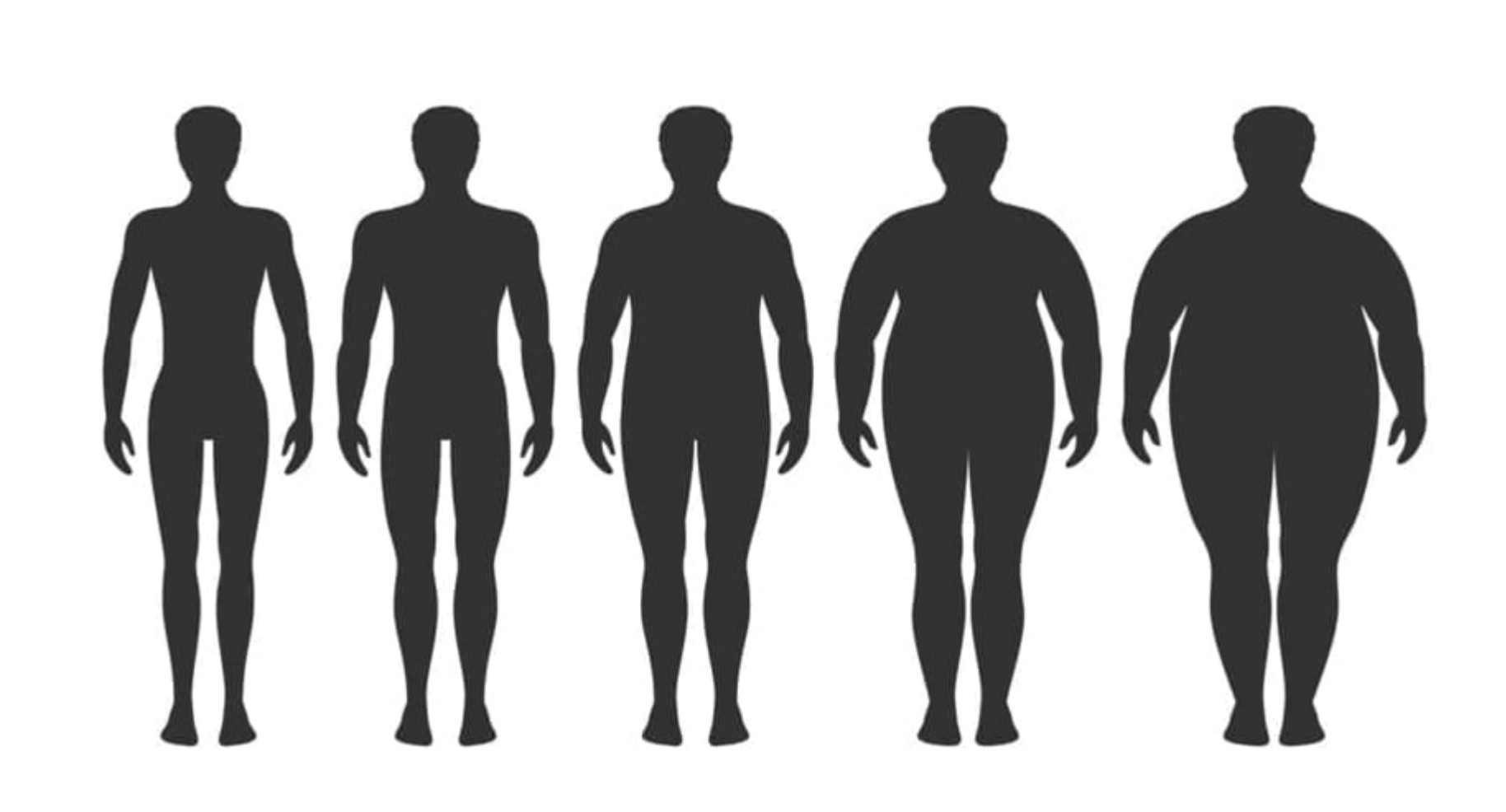

**4.**

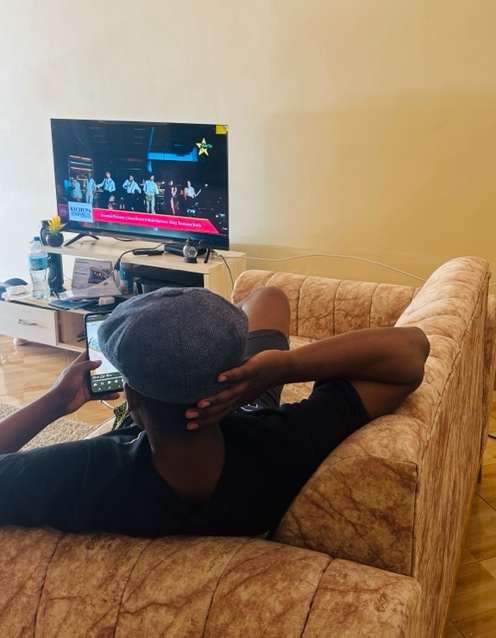

**5.**

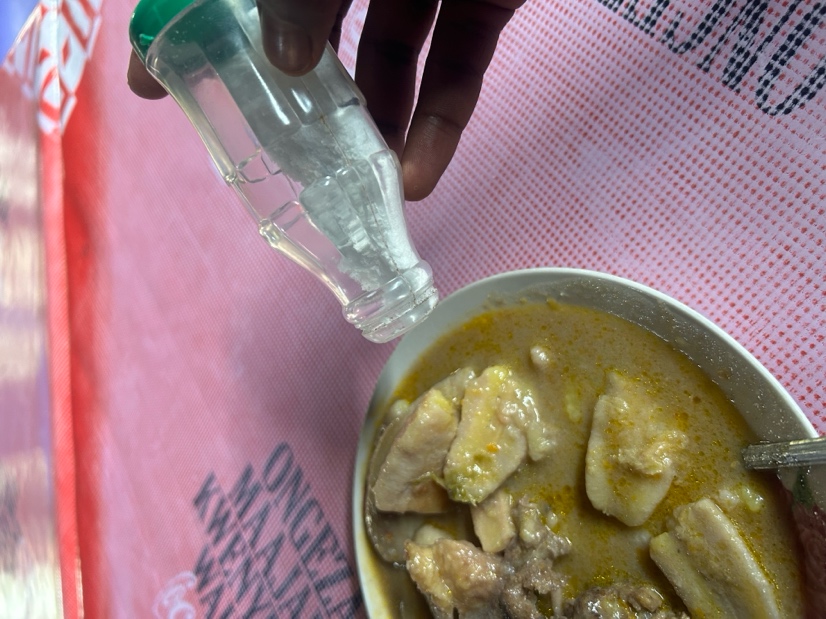

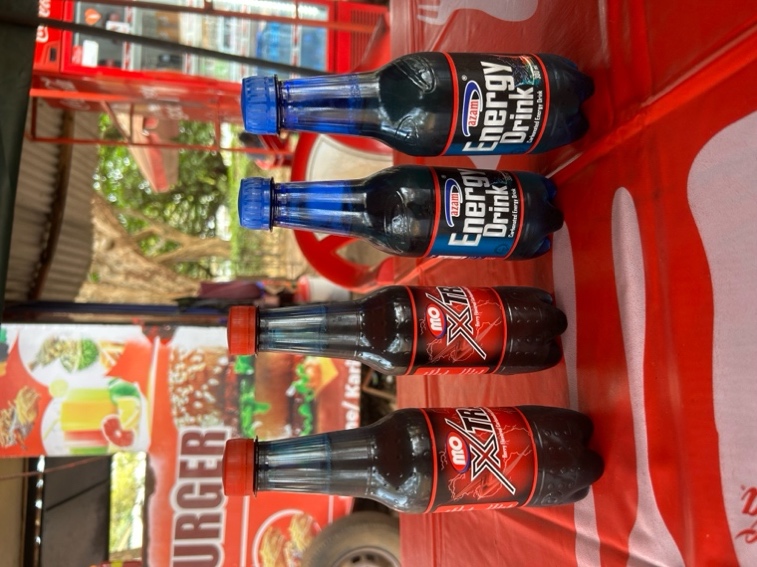
**6.**

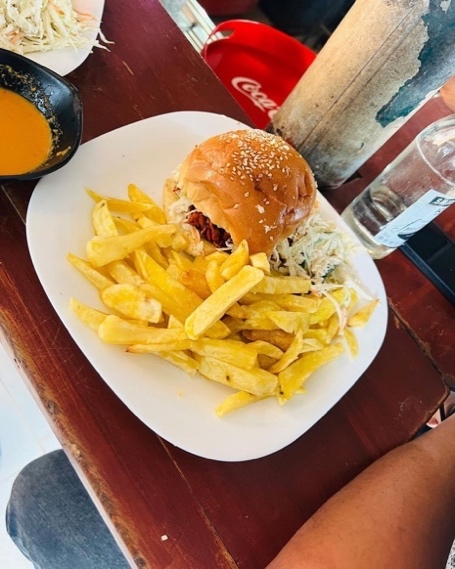

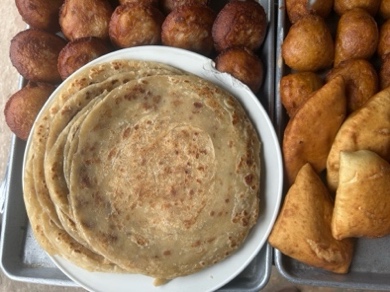

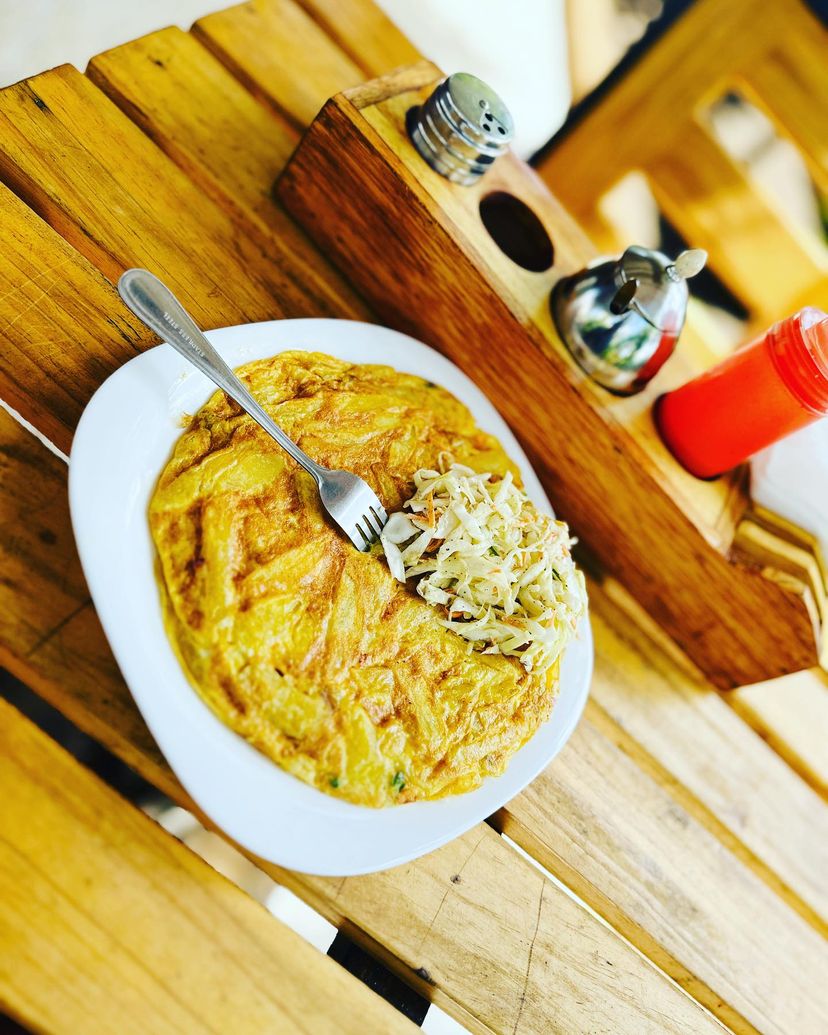

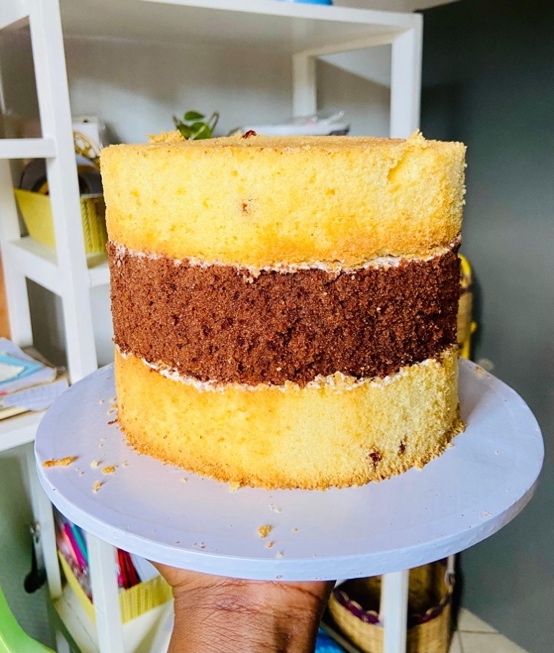

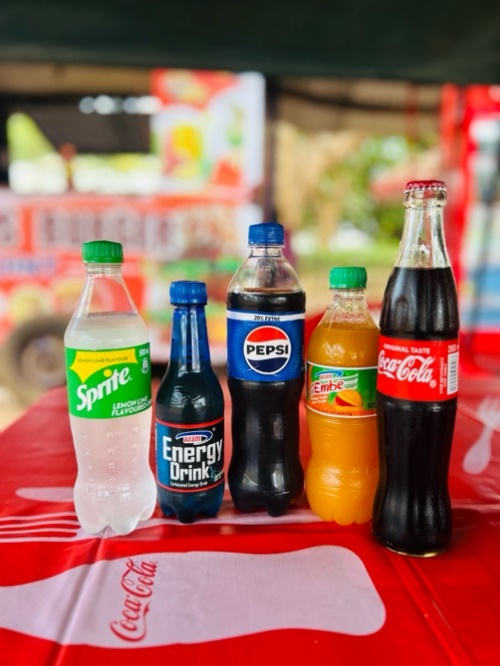

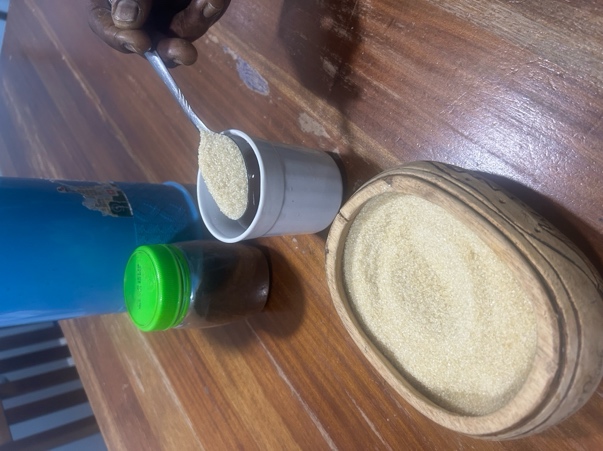

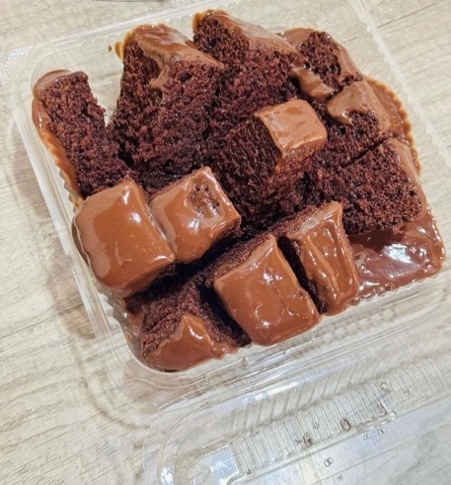

**7.**

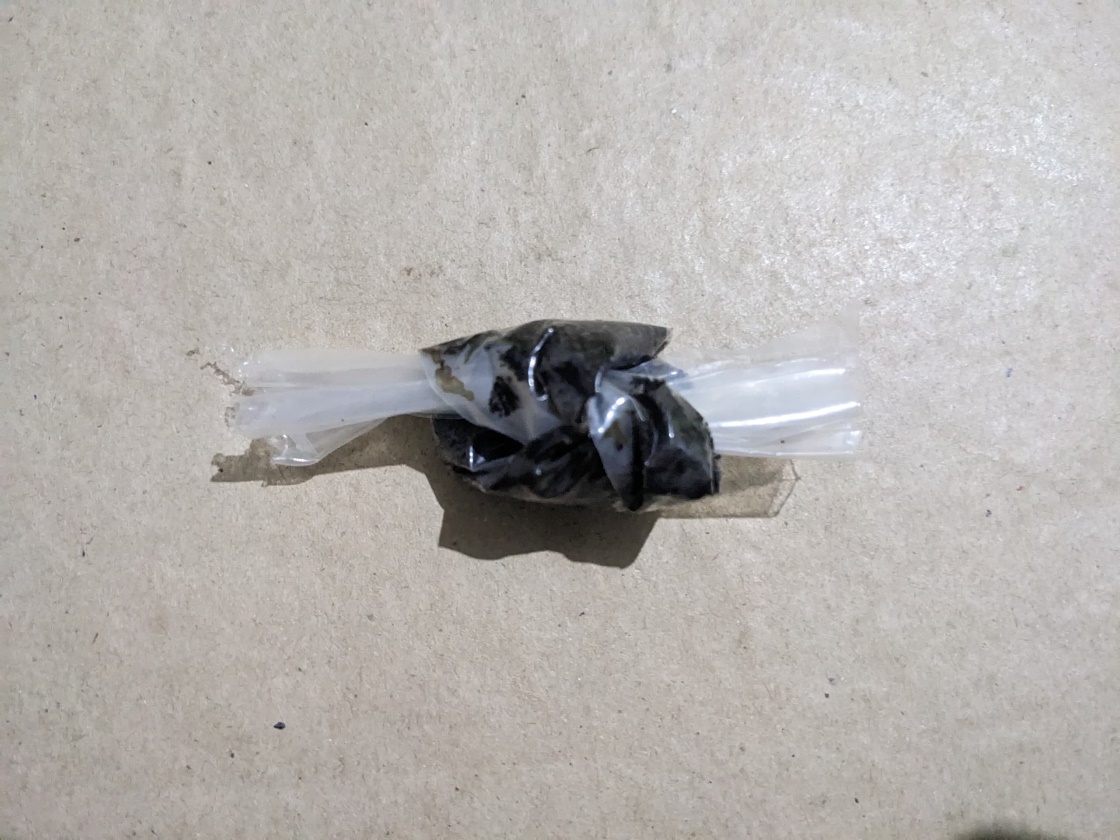

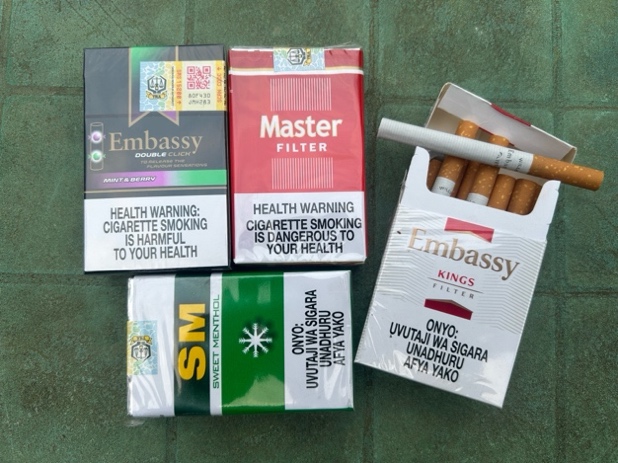

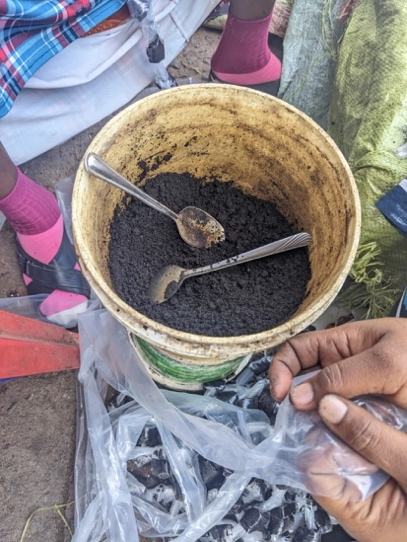

**8.**

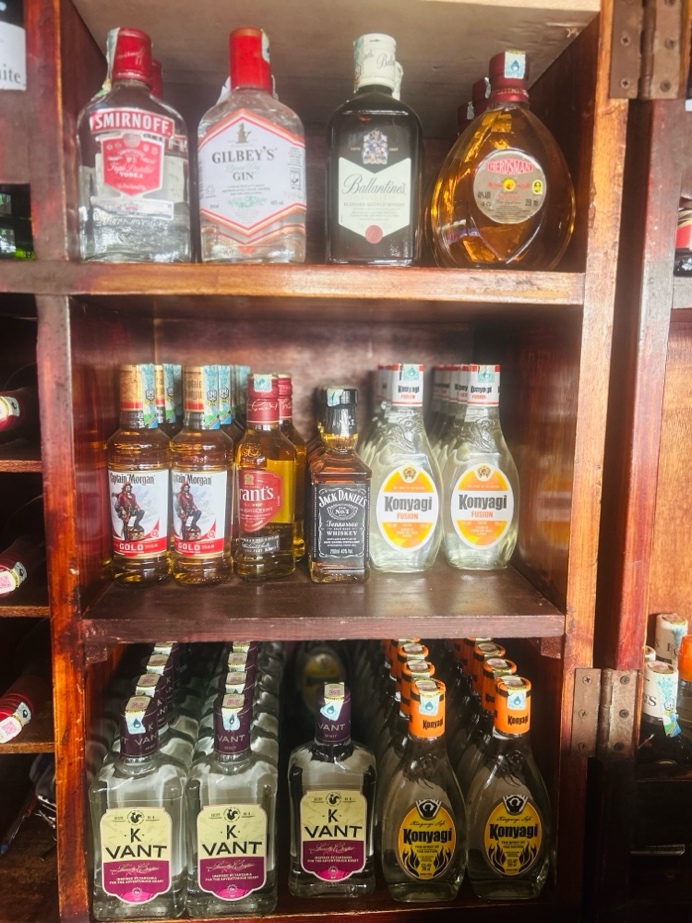

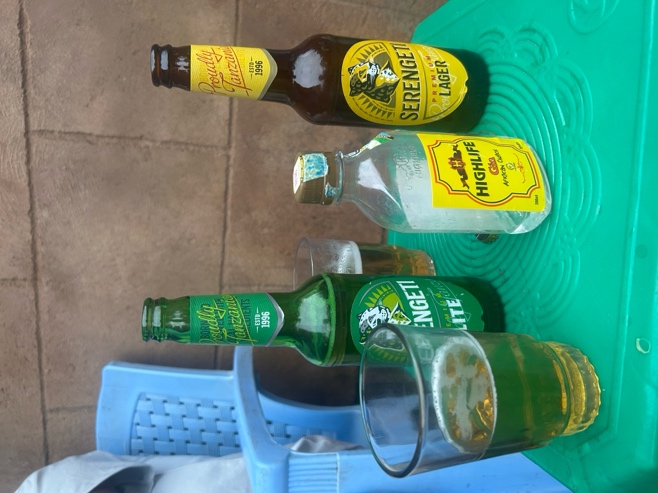

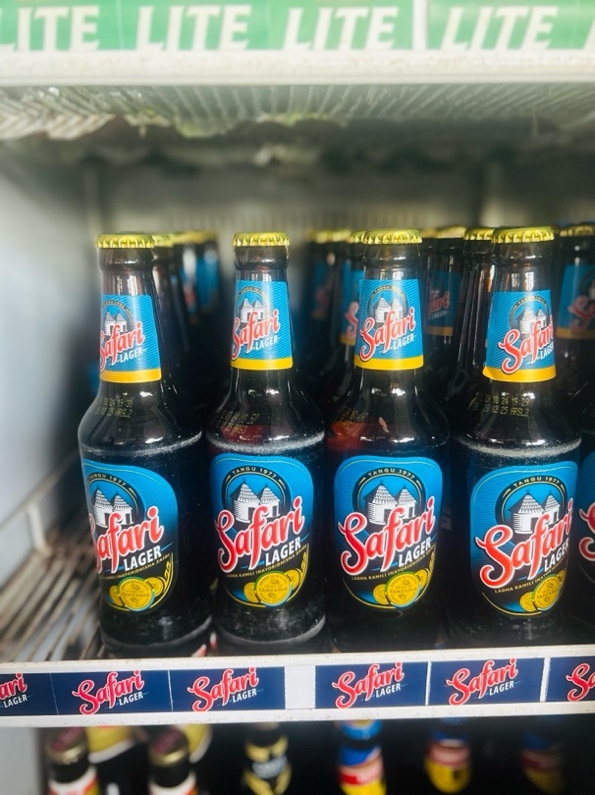

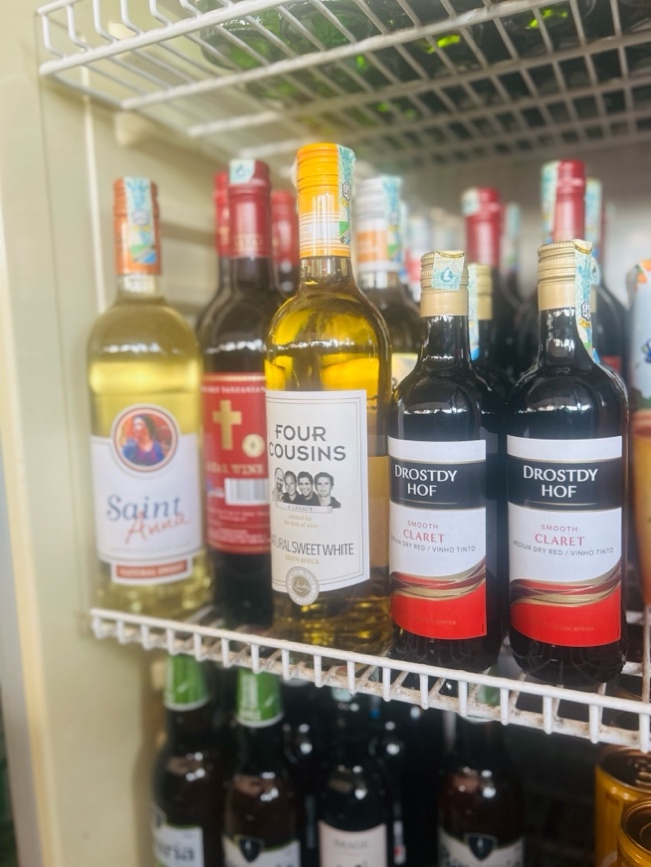

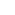

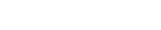

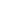

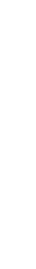

**11.**

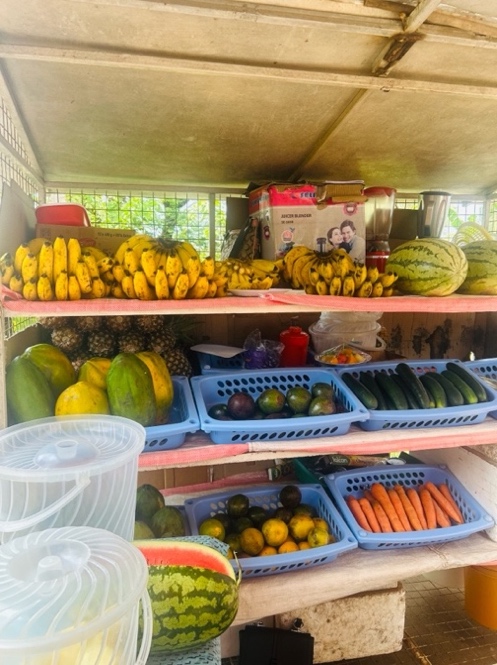

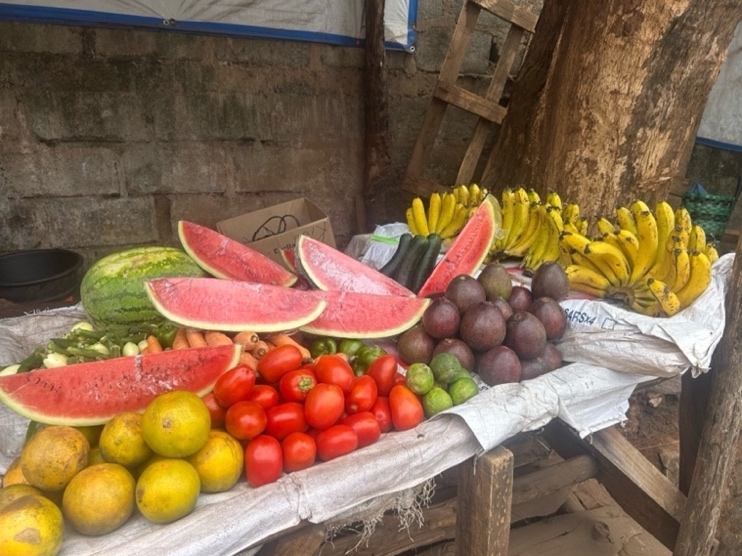

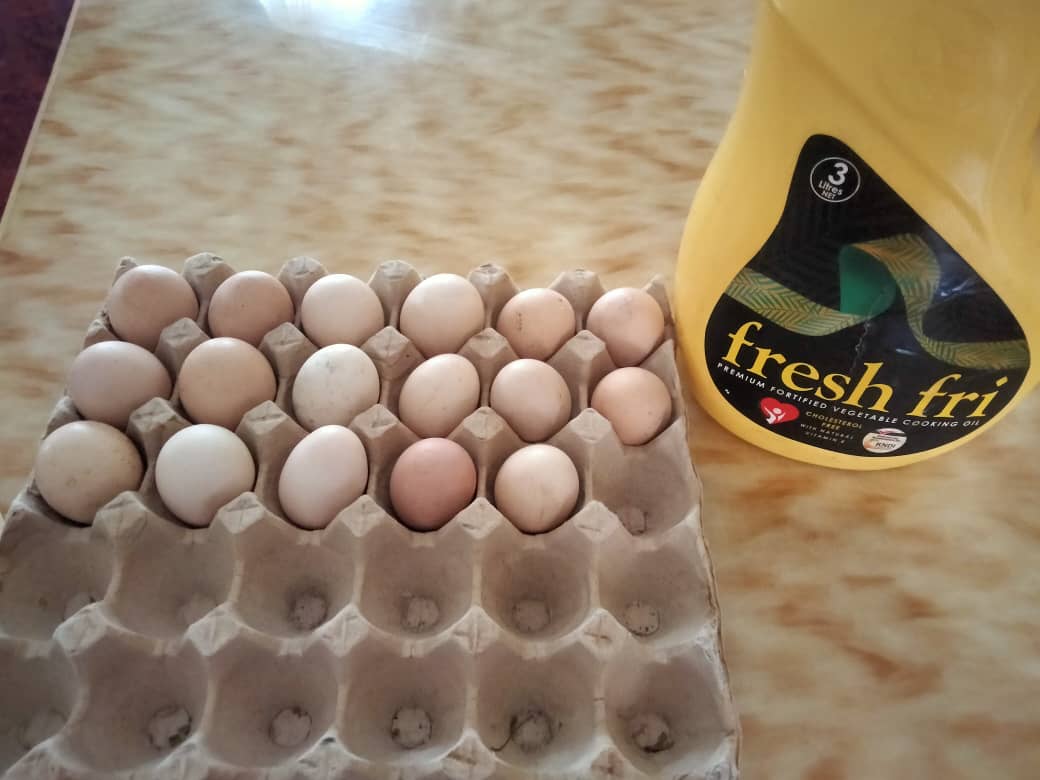

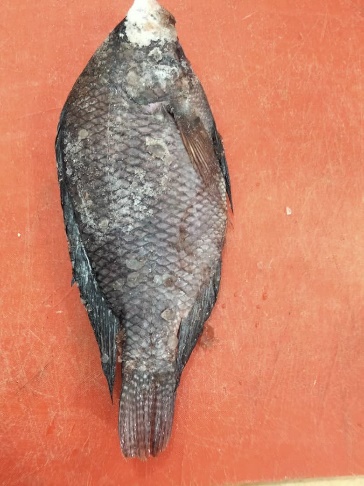

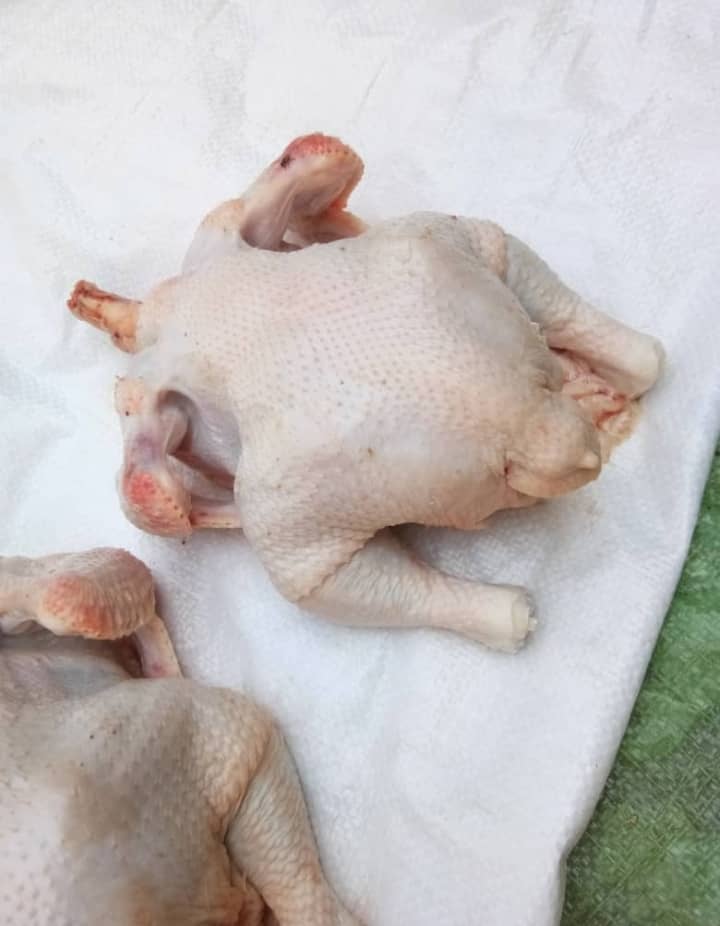

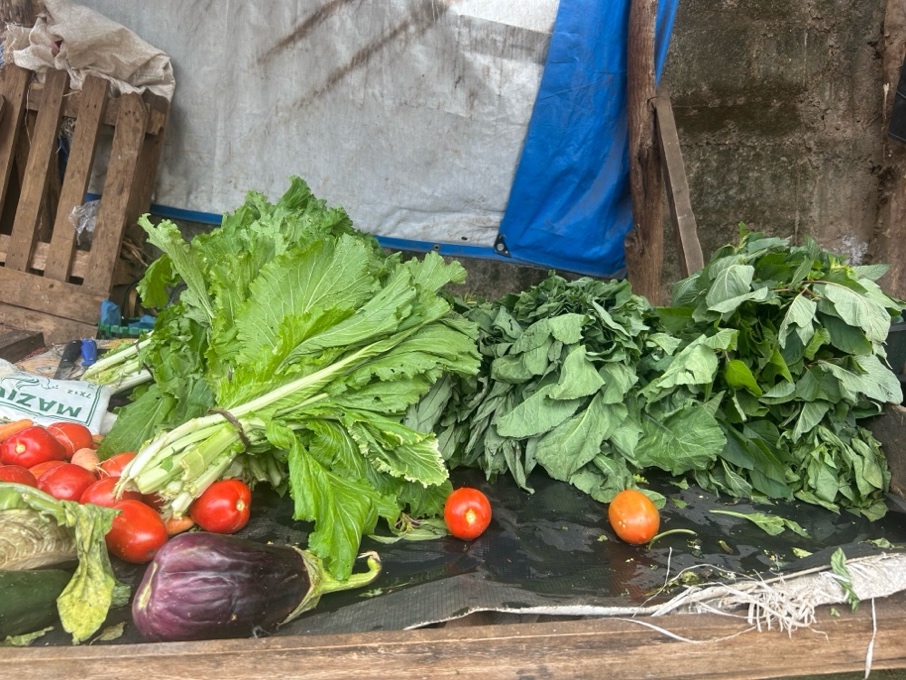

**13.**

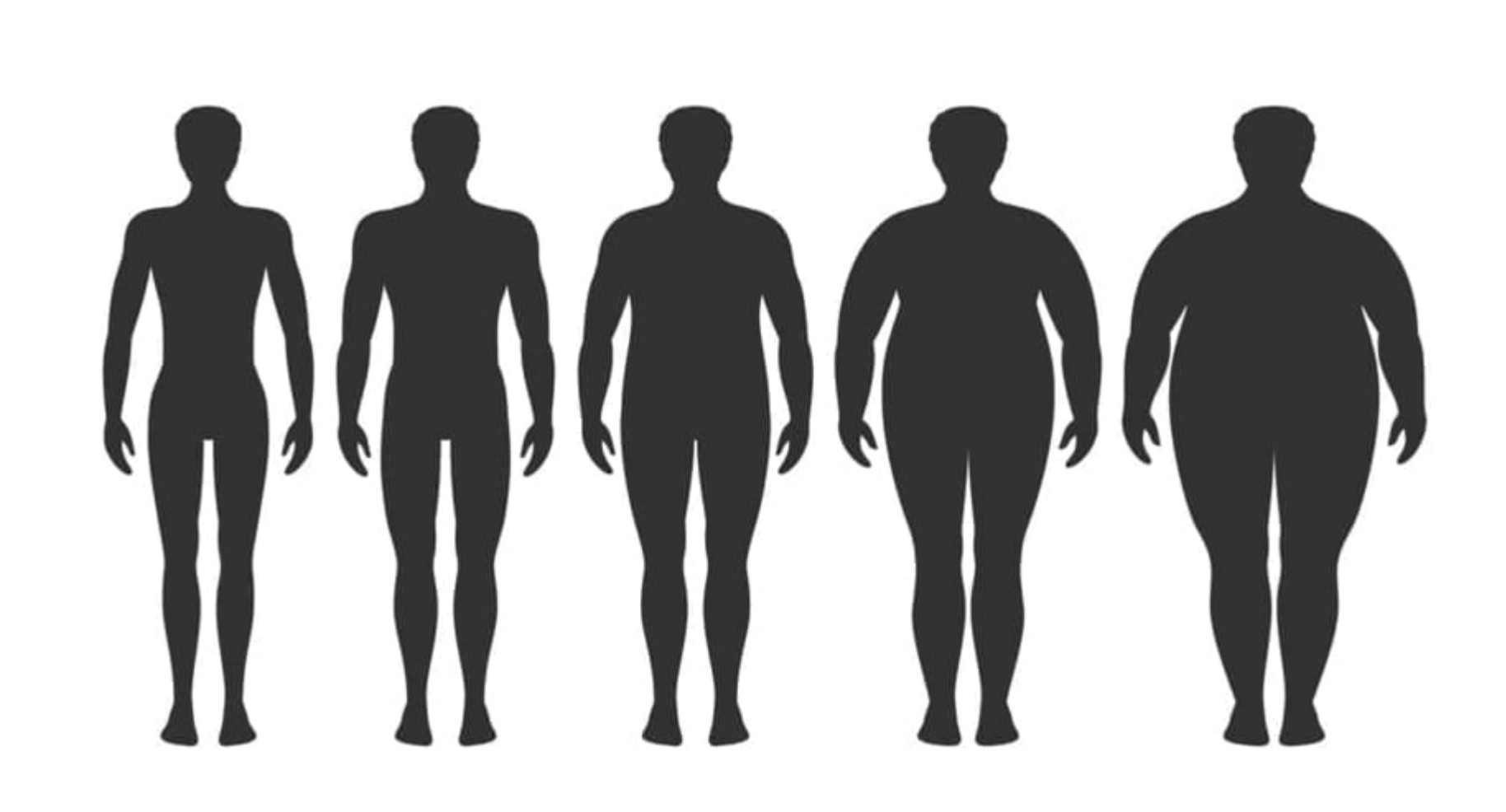

**15.**

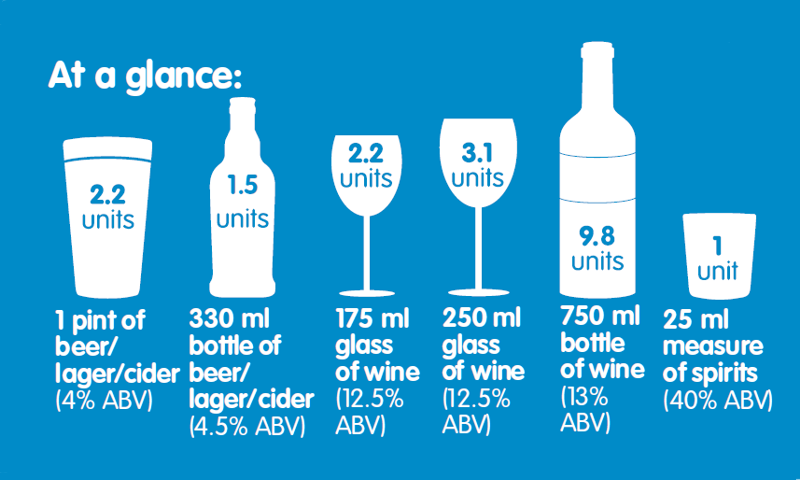
